# Targeted memory reactivation during sleep modulates spindle and slow wave density, but not motor memory consolidation, in Parkinson’s disease

**DOI:** 10.64898/2026.04.22.26351552

**Authors:** Letizia Micca, Genevieve Albouy, Bradley R King, Alice Nieuwboer, Wim Vandenberghe, Pascal Borzée, Bertien Buyse, Dries Testelmans, Judith Nicolas, Moran Gilat

## Abstract

Motor memory retention is impaired in Parkinson’s disease (PD), affecting long-term rehabilitation outcomes. It appears that NREM sleep could be beneficial for consolidation processes in PD, and could be leveraged with non-invasive sleep interventions. This study examined the effect of auditory targeted memory reactivation (TMR) during NREM sleep on the retention of a motor sequence learning finger tapping task in 20 PD and 20 healthy older adults (HOA). TMR was applied during a 2- hour nap and its effect on motor retention was tested post-nap, after 24-hours and with a dual-task. The impact of TMR on sleep electrophysiology was also evaluated. Results showed no effect of TMR on motor retention or dual-tasking, with no difference between the groups. However, the TMR intervention did increase slow-wave density and decreased spindle density in both groups, and slow-wave amplitude during the presentation of the auditory cues was positively associated with performance in HOA. In conclusion, TMR applied during a 2-hour nap did not enhance motor retention, but the changes in sleep physiological features could be linked to a possible underlying effect on memory processing that warrants further investigation.

## Introduction

Parkinson disease (PD) is a complex neurological disorder, characterised by the progressive loss of dopaminergic neurons in the substantia nigra pars compacta and consequent dysfunction of the striatum (Morrish et al., 1996). The pathophysiological alterations observed in PD lead to several motor and non-motor symptoms that require multidisciplinary management (Jankovic & Tan, 2020). The therapy for PD often involves rehabilitation modalities that leverage motor acquisition and consolidation mechanisms, aimed to restore impaired daily-life functions (Nieuwboer, Rochester, Müncks, et al., 2009). These processes of motor learning have been linked to striatal function, and can thus become progressively impaired in PD (Nieuwboer, Rochester, Müncks, et al., 2009).

Specifically, skill acquisition, defined as the online process occurring during task practice (Karni et al., 1995, 1998), has been related to anterior striatal functioning, which is relatively less affected in the early stages of PD (Wu et al., 2015). With adequate amounts of practice, initial skill acquisition in PD can therefore be comparable to that of healthy older adults (HOA). Conversely, consolidation and automaticity are particularly associated to posterior striatal functioning (Doyon et al., 2009; Puttemans et al., 2005), which is most affected in PD. Consolidation refers to the post-learning offline process of memory strengthening happening without practice, assessed with a retention test (Brashers-Krug et al., 1996; Karni et al., 1998; Robertson et al., 2004). Automaticity refers to the reduced need of focused attention on the task, allowing for performance of a secondary dual-task (Fitts & Posner, 1967). Retention and automaticity deficits can therefore already be present early in the disease (Nieuwboer, Rochester, Müncks, et al., 2009; Nieuwhof et al., 2017).

Interestingly, the hippocampus is also involved in motor memory consolidation (Albouy et al., 2008, 2013), particularly during non-rapid eye movement (NREM) sleep (King et al., 2017). This brain structure seems relatively spared in the early stages of PD (Nurmi et al., 2001), and its role in sleep-dependent memory consolidation networks could be leveraged with sleep interventions promoting compensatory long-term neuroplastic changes, as previously studied in healthy young and ageing populations (Muehlroth et al., 2019; Staresina, 2024). Such approaches could partially bypass the dysfunctional striatum and could represent a valuable add-on therapy to rehabilitation in PD.

Sleep-dependent memory consolidation is thought to rely on spontaneous reactivation of the newly formed neuronal patterns associated to the memory traces encoded during training. Specifically, the electrophysiological activity during NREM sleep stages seems to promote these reactivations (Born & Wilhelm, 2012) thanks to the temporal coordination of hippocampal sharp wave-ripples (80–100Hz frequency band) (Axmacher et al., 2008), thalamic spindles (12–16Hz, sigma band) (Berry et al., 2018; Iber, 2007) and cortical slow waves (0.5–2 Hz, delta band) (Ngo et al., 2013). While these models were initially developed for declarative memories, they are thought to also apply to hippocampal-mediated motor memory traces (Albouy et al., 2013).

Sleep-dependent consolidation is reduced with ageing (S. M. Fogel et al., 2014; King et al., 2013; Spencer et al., 2007; Wilson et al., 2012), with consequent electrophysiological alterations of slow waves (Mander et al., 2013). Despite the age-related deterioration of consolidation mechanisms, the implementation of additional practice or the choice of specific tasks adapted to the population’s age can still partially support these processes (Fitzroy et al., 2021; Gudberg et al., 2015; King et al., 2016). In PD, sleep-dependent memory consolidation of a motor sequence learning (MSL) task is similar to HOA, when measured immediately after a period of sleep (Dan et al., 2015; Lanir-Azaria et al., 2024; Micca et al., 2025; Terpening et al., 2013). This suggests that while people with PD in the early stages can have issues with long-term retention and automaticity, the consolidation processes happening during sleep may be similar to those observed in HOA. Interestingly, electrophysiological markers of consolidation, specifically slow wave characteristics, appear positively associated to behavioural performance improvements in people with PD after a 2-hour nap (Micca et al., 2025), suggesting that sleep interventions targeting slow waves could allow for further performance benefits and potentially minimize the retention deficit observed in PD.

In this context, targeted memory reactivation (TMR) during sleep is a promising non-invasive intervention whereby a sensory cue is associated with to-be-learned material during initial acquisition, and replayed during subsequent sleep (Temudo & Albouy, 2024). Its application has been investigated with different stimulation modalities (e.g., auditory, olfactory, tactile) (Hu et al., 2020). While the use of olfactory or tactile cues may prove less feasible in PD due to hyposmia (Jankovic & Tan, 2020) and somatosensory deficits (Schindlbeck et al., 2016), the use of auditory cues for TMR could represent a beneficial modality in this population. Moreover, slow waves, and specifically k-complexes, can be evoked with auditory stimulation (Bellesi et al., 2014). TMR could thus promote the modulation of slow-wave activity during sleep, and to a broader extent, the consolidation of memory content associated to the stimuli. The application of auditory TMR during NREM2 and NREM3 sleep has been studied in the healthy young and older populations as a tool to improve memory retention (Carbone & Diekelmann, 2024; Hu et al., 2020b). Young adults consistently showed a behavioural benefit of TMR on the consolidation of motor sequence memories (Antony et al., 2012; Cousins et al., 2016; Nicolas et al., 2022, 2025; Schönauer et al., 2014), with an enhancement of activity of slow waves and of their coupling with spindles when TMR was applied (Nicolas et al., 2022).

The effect of TMR on motor performance in older adults is less consistent. While work by Johnson et al. indicated improvements in sensorimotor performance after a post-learning nap with TMR in HOA (Johnson et al., 2020), compared to an equivalent time without the intervention, the study by Nicolas and colleagues showed no such benefit on a MSL task (Nicolas et al., 2024). However, auditory stimulation did result in modulations of slow wave-spindle coupling irrespective of whether the stimulus was associated to memory content or whether it was a sham stimulus (Nicolas et al., 2024). Hence, auditory stimulation might contribute to the modulation of memory consolidation processes during sleep in the ageing brain, though it could act in a non-specific manner, irrespective of whether the cue presented was previously paired to a motor task or not. To date, the evidence of the effects of sleep interventions and the associations between sleep electrophysiology and motor memory consolidation in PD remains largely understudied, and results are often controversial. Testing the effects of auditory TMR could shed light on the potential of sleep interventions in PD.

Accordingly, the aim of this study was to evaluate for the first time the effects of auditory TMR applied during a 2-hour post-learning nap on the consolidation in PD. Specifically, we set out to study the behavioural and electrophysiological changes generated by this intervention during sleep in PD and in HOA, finding that auditory stimulation, used in a TMR paradigm, induces electrophysiological changes of NREM2 and NREM3 sleep characteristics in both groups. We could not demonstrate a TMR-specific effect on immediate post-intervention and 24-hour retention of the motor task in PD. Finally, despite prior associations found between slow waves and retention performance after a 2-hour nap in people with PD (Micca et al., 2025), we did not find an association between sleep electrophysiological metrics measured when the auditory stimulation was applied, and performance at post-nap retention in patients. However, this was evident in the HOA group, with positive associations between performance and slow-wave amplitude.

## Results

### Participants’ Characteristics

Participants’ demographic and clinical characteristics are listed in Table 1. The groups were matched for demographic characteristics, though the PD group self-reported worse sleep. The PD cohort had mild-to-moderate disease severity (Hoehn and Yahr stages II-III). Moreover, five people with PD and one HOA reported active use of benzodiazepines and/or antidepressants during the study.

**Table 1.**
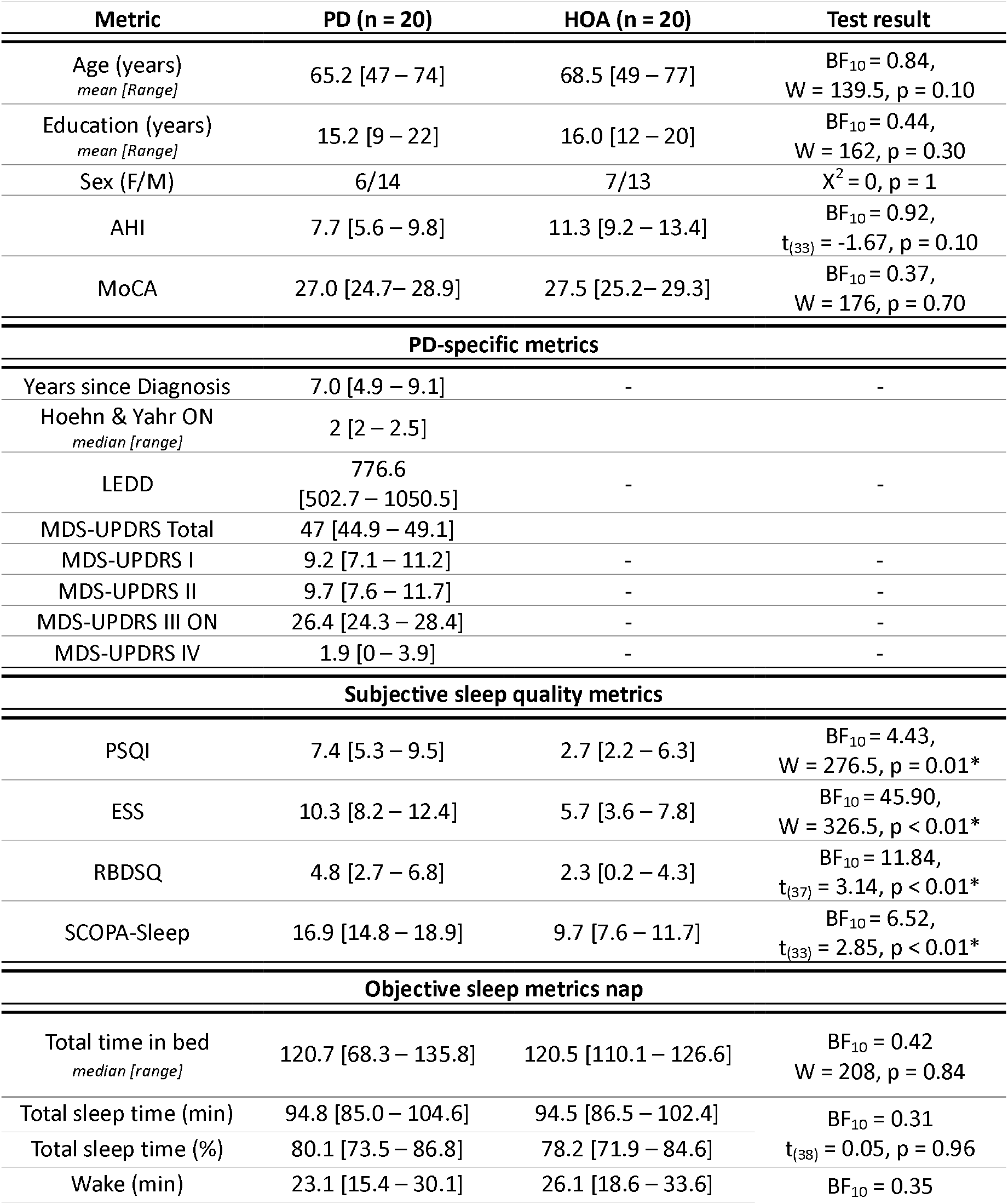
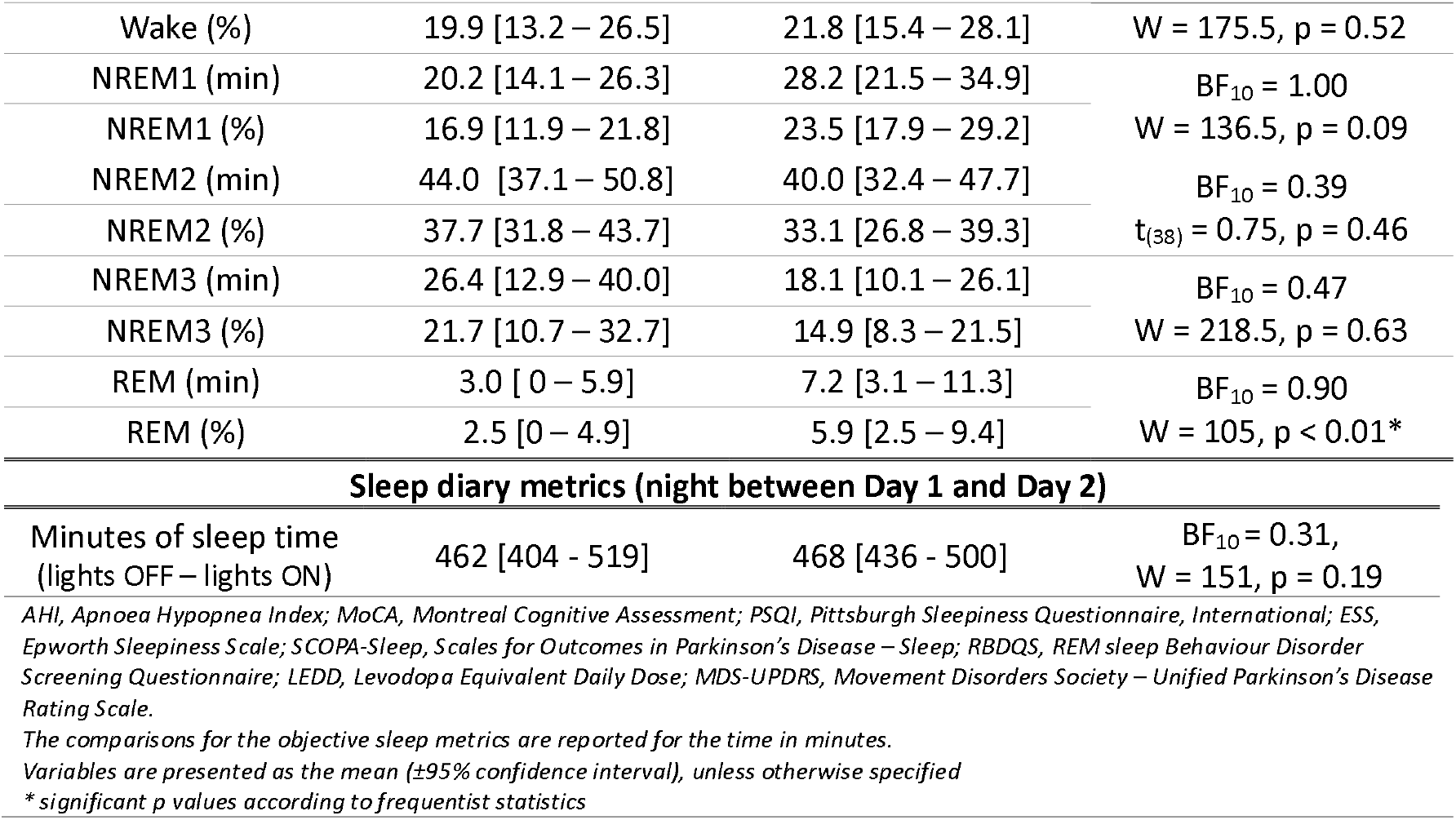
Demographic characteristics of the experiment cohort.

### Behavioural Performance Does not Benefit from TMR in PD and HOA

Performance was measured during three experimental sessions (i.e., pre-nap, post-nap and 24-hour retention) (see Figure 1.A). The main outcomes were reaction time and accuracy of two serial reaction time task sequences (SRTT), each cued with a distinct melody of auditory tones. TMR was applied in a within-subject fashion, whereby only one of the two melodies was randomised to be reactivated during the 2-hour nap intervention for each participant. Given the explorative nature of this study, we opted for reporting Bayesian statistics, which provide an indication of the effects in case of non-significant findings. Firstly, we report the results regarding initial motor acquisition of the two sequences. Secondly, we describe the effects of TMR on the motor outcomes, and finally we report the effects of the TMR intervention on sleep electrophysiology, comparing between periods with and without the application of TMR in the same subject (Figure 1.B).

**Figure 1.**
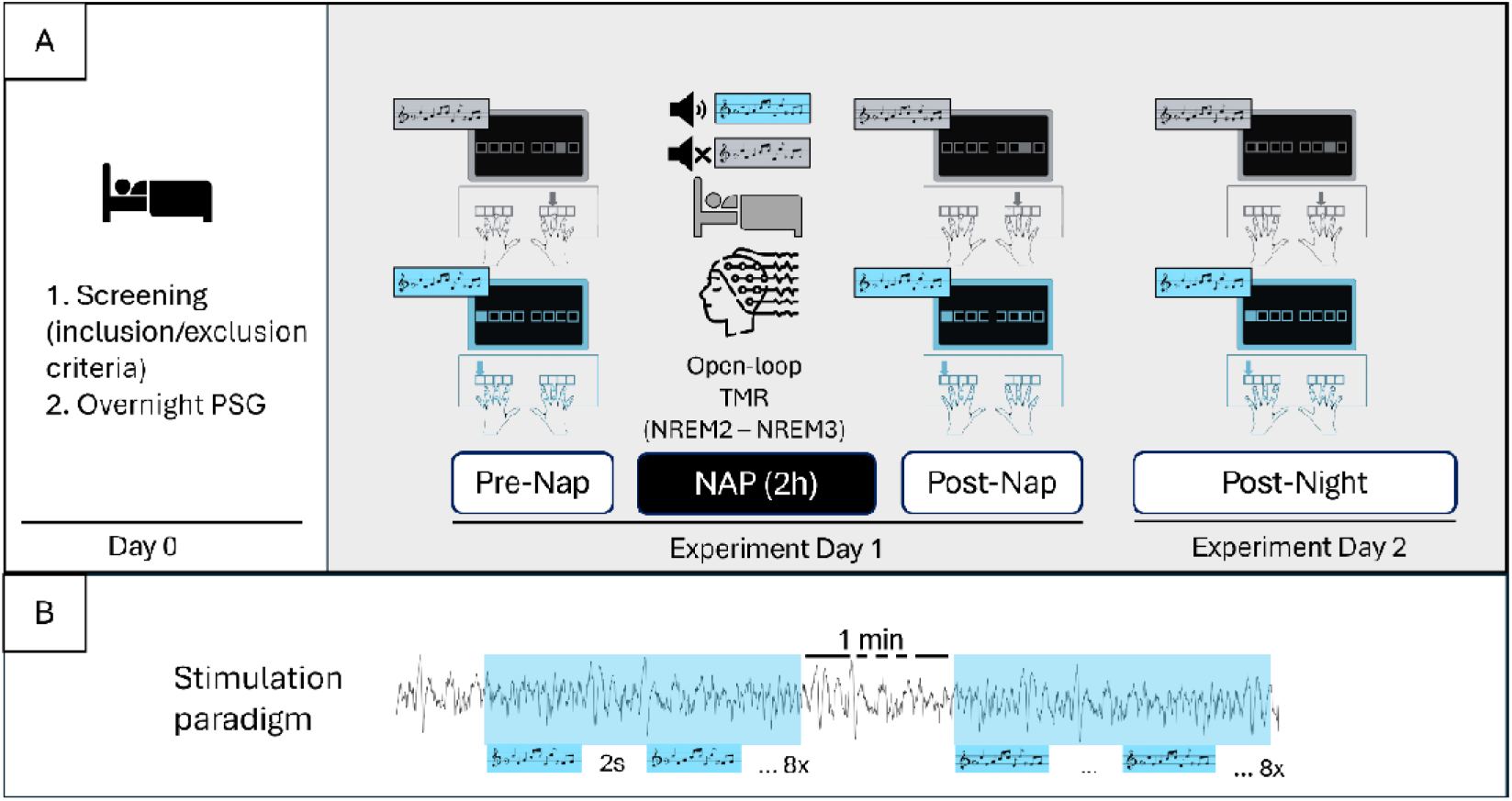
Experimental design. (A) After a screening night and evaluation of inclusion and exclusion criteria, 20 people with PD and 20 HOA were invited for the experiment, consisting of an pre-nap session, during which they practiced two moor sequences in a SRTT task, each cued with a distinct sequence of sounds. This pre-nap initial learning session was followed by a 2-hour nap with TMR, during which only one of the auditory sequences was randomised to be replayed. Subsequently, participants underwent a post-nap session, during which their motor performance of the two sequences was re-assessed, both in single-and dual-task, to test automaticity. After a night of sleep at home, they were finally invited for a 24-hour retention test, during which performance of the motor sequences was tested once again, both in single- and dual-task. (B) The auditory stimulation during sleep was applied during online scored epochs of NREM2-NREM3 sleep. The sequence of auditory tones was replayed 8 consecutive times, with an intersequence latency time of 2s. After the 8 repetitions, there followed 1 minute silence. Stimulation was manually stopped in case of arousal or change of sleep stage. PSG = polysomnography; TMR: targeted memory reactivation.

#### Acquisition

While the two sequences were learnt similarly, differences between groups were found for the initial motor acquisition rate measured with reaction time (RT) (see Pre-nap panels in Figure 2A-B), but not with accuracy (see Pre-nap panels in Figure 3A-B, and the **Supplementary Material, Initial Encoding**). Specifically, motor acquisition of the two sequences was greater in the HOA group as compared to the PD group, when measured with RT. Importantly, the comparisons between the sequential SRTT and the random SRTT across sessions suggest that the performance improvements were not exclusively related to motor execution (see **Supplementary Material, Motor Learning**).

**Figure 2.**
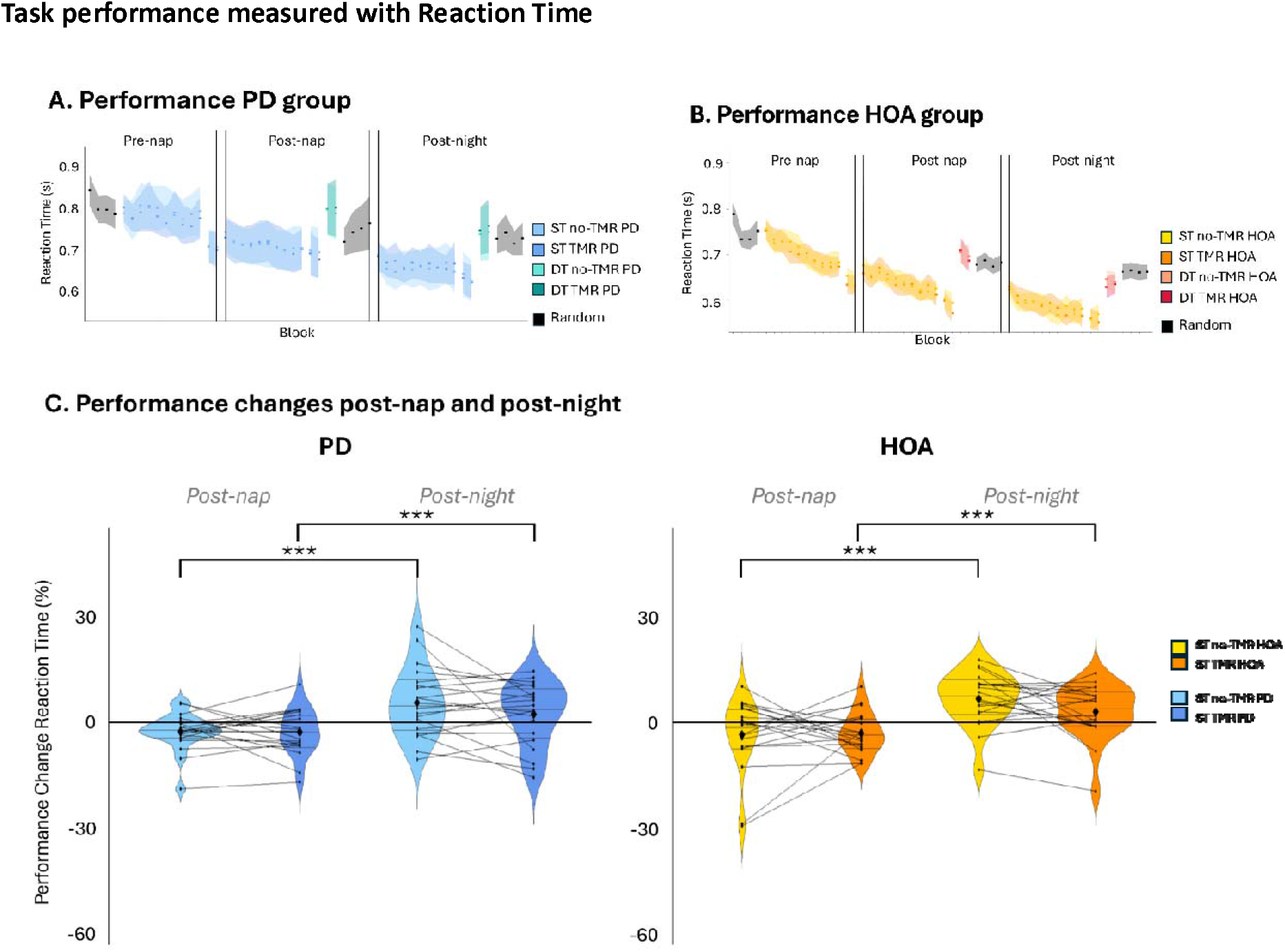
Performance measured with reaction time. (A) Performance across sessions of the PD group and of the (B) HOA group. The dots represent the mean of the median values of each subject, the ribbon indicates the standard error. ST: Single-task; DT: Dual-task. (C) Single-task performance changes measured with reaction time (RT), where higher values indicate better performance. Violin plot: mean (diamond), median (central horizontal bar), and 25^th^ (lower bar) and 75^th^(higher bar) percentiles. *** Decisive evidence for a difference

**Figure 3.**
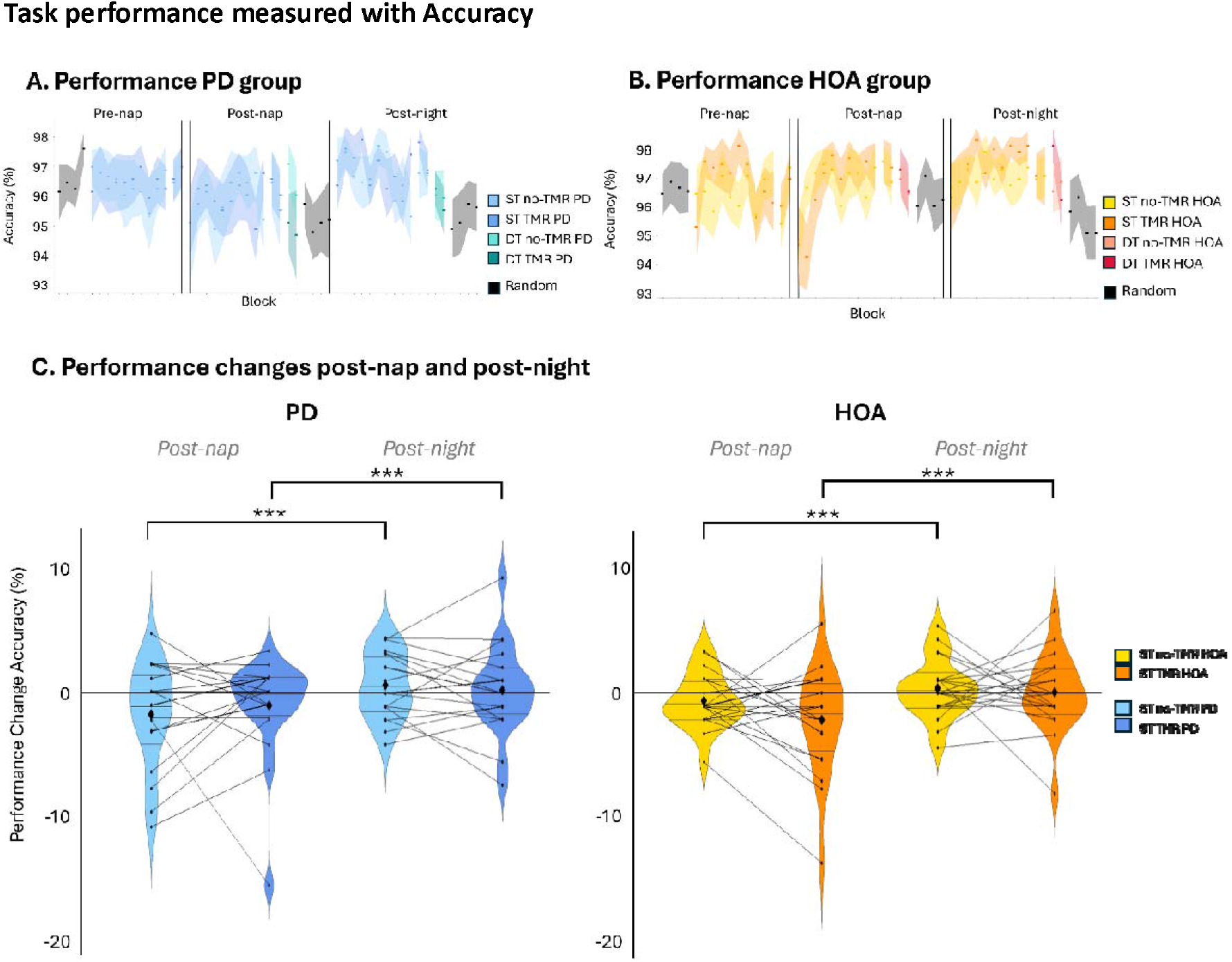
Performance measured with accuracy. (A) Performance across sessions of the PD group and of the (B) HOA group. The dots represent the mean of the median values of each subject, the ribbon indicates the standard error. ST: Single-task; DT: Dual-task. (C) Single-task performance changes measured with reaction time (RT), where higher values indicate better performance. Violin plot: mean (diamond), median (central horizontal bar), and 25^th^(lower bar) and 75^th^ (higher bar) percentiles. *** Decisive evidence for a difference

#### Consolidation

The relative changes between performance at the end of initial acquisition (last four blocks of retest pre-nap) and the beginning of the post-nap (first four blocks of practice post-nap) and post-night (first four blocks of practice post-night) sessions were used as a proxy of consolidation for both RT and accuracy. Bayesian ANOVAs, complemented with frequentist ANOVAs including group (PD/HOA), condition (reactivated/non-reactivated), time (post-nap/post-night) and their interactions as fixed effects, and subject as random effect were used to study immediate consolidation and 24-hour retention.

##### Reaction time

Decisive evidence for an effect of time was found (BF_10_ > 100, F_(1,38)_ = 35.07, p < 0.01, ges = 0.17, post-hoc: BF_10_ > 100, V = 396, p < 0.01, r_rb_ = 0.45, Figure 2), suggestive of a performance improvement of both sequences across sessions. However, such RT improvement did not differ between groups, nor between reactivated and non-reactivated sequences, as indicated by the weak to moderate evidence for no other main effects or interactions (group: BF_10_ = 0.24, F_(1,38)_ = 0.06, p = 0.81, ges < 0.01; condition: BF_10_ = 0.40, F_(1,38)_ = 1.78, p = 0.19, ges = 0.01; group by condition interaction: BF_10_ = 0.24, F_(1,38)_ = 0.0008, p = 0.98, ges < 0.01; group by time interaction: BF_10_ = 0.25, F_(1,38)_ = 0.21, p = 0.65, ges < 0.01; condition by time interaction: BF_10_ = 0.60, F_(1,38)_ = 6.90, p = 0.01, ges = 0.01; group by condition by time interaction: BF_10_ = 0.30, F_(1,38)_ = 0.14, p = 0.71, ges < 0.01).

##### Accuracy

Similarly, performance changes measured with accuracy showed decisive evidence for an effect of time (BF_10_ > 100, F_(1,38)_ = 17.04, p < 0.01, ges = 0.07; post-hoc: BF_10_ > 100, V = 436.5, p < 0.01, r_rb_ = 0.22), which were not different between groups or reactivated and non-reactivated sequences (group: BF_10_ = 0.25, F_(1,38)_ = 0.01, p = 0.92, ges < 0.01; condition: BF_10_ = 0.23, F_(1,38)_ = 0.50, p = 0.48, ges < 0.01; group by condition: BF_10_ = 0.38, F_(1,38)_ = 0.88, p = 0.35, ges < 0.01; group by time: BF_10_ = 0.24, F_(1,38)_ = 0.12, p = 0.73, ges < 0.01; condition by time: BF_10_ = 0.23, F_(1,38)_ = 0.005, p = 0.94, ges < 0.01; group by condition by time: BF_10_ = 0.53, F_(1,38)_ = 2.27, p = 0.14, ges < 0.01) (Figure 3).

Overall, performance of RT and accuracy improved over time, but the consolidation of the sequence reactivated with TMR was not increased as compared to the non-reactivated sequence, nor did we find differences in performance between groups.

#### Dual-task Results

A working memory dual-task was performed (i.e. counting squares that appeared in a different colour). The cost was computed as the relative difference between performance of the four blocks of dual-task and the four blocks of test single-task post-nap and post-night separately. Bayesian ANOVAs including group, condition, time and their interactions as fixed effects, and subject as random effect were used for the analyses. Unexpectedly, findings suggest that TMR did not affect dual-tasking, neither immediately after the intervention, nor at 24-hour retention in both groups. The specific results for RT and accuracy are reported in **Supplementary Material, Dual-task Results**.

### TMR Modulates Spindle and Slow Wave Density, but not Amplitude

Spindle and slow wave characteristics were extracted from the 2-hour nap intervention and compared between periods with and without the application of auditory stimuli (Figure 1.B), to allow for the investigation of the modulatory effects of this intervention. The amount of stimulations sent in each sleep stage did not differ between groups (see **Supplementary Material, Stimulations per Sleep Stage**).

#### Density

##### Spindles

When comparing spindle density during periods with TMR and silent periods, strong evidence for an effect of the condition was found, suggesting a reduced amount of spindles in the TMR periods (BF_10_ = 13.93, F_(1,34)_ = 10.49, p < 0.01, ges = 0.03, post-hoc comparison: BF_10_ = 14.98, t_(35)_ = 3.29, 95% CI [0.32 – 1.36], p < 0.01, Cohen’s d = 0.32, Figure 4Figure 4.A). However, there was weak evidence for no difference between people with PD and HOA (BF_10_ = 0.56, F_(1,34)_ = 0.40, p = 0.53, ges = 0.01) and for no interaction effect of condition and population (BF_10_ = 0.33, F_(1,34)_ = 0.004, p = 0.95, ges < 0.01).

**Figure 4.**
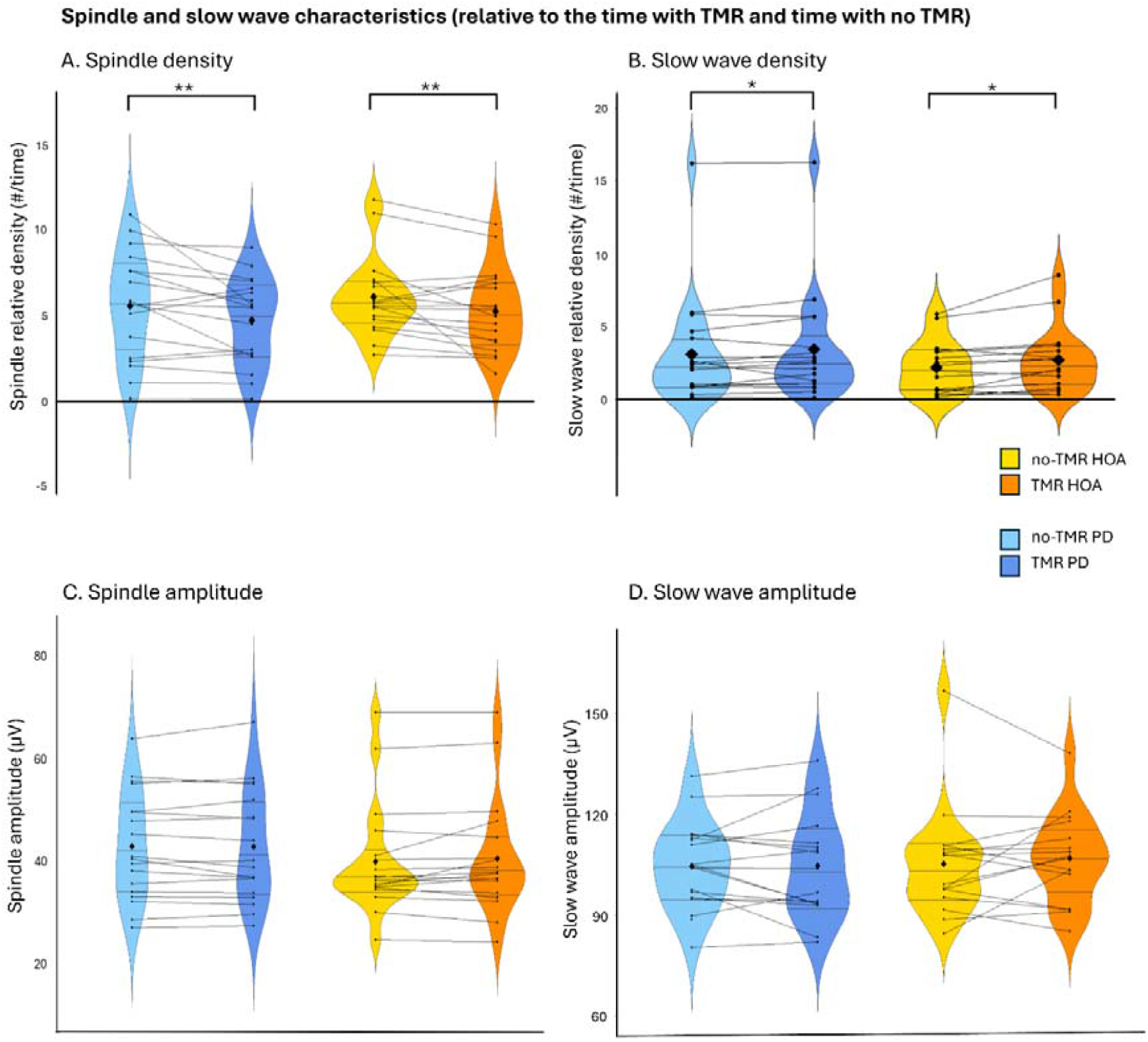
Density of (A) spindles and (B) slow waves, measured relative to the amount of NREM2 and NREM3 sleep spent with auditory stimulation, and time spent with no auditory stimulation. Amplitude of (C) spindles and (D) slow waves during the auditory stimulation and silent periods. Violin plot: mean (diamond), median (central horizontal bar), and 25^th^ (lower bar) and 75^th^ (higher bar) percentiles. Spindle data: 17 people with PD and 17 HOA in the TMR periods, and for 17 people with PD and 17 HOA in the silence periods. Slow wave data: 16 people with PD and 17 HOA in the TMR periods, and for 17 people with PD and 17 HOA in the silence periods. ** strong evidence in favour of a difference; * moderate evidence in favour of a difference.

##### Slow waves

Conversely, slow wave density increased during the periods when auditory stimulation was applied, supported by a moderate effect of the condition (BF_10_ = 3.24, F_(1,31)_ = 6.87, p = 0.01, ges < 0.01, post-hoc comparison: BF_10_ = 3.15, V = 134, p < 0.01, r_rb_= 0.07, Figure 4.B). We found weak evidence for no effect of group (BF_10_ = 0.76, F_(1,31)_ = 0.70, p = 0.41, ges = 0.02) or group by condition interaction (BF_10_ = 0.59, F_(1,31)_ = 2.06, p = 0.16, ges < 0.01).

These results suggest that spindle and slow wave density are modulated in different ways by the auditory stimulation, whereby spindle density is reduced and slow wave density is increased both in people with PD and HOA.

#### Amplitude

TMR did not have a modulatory effect on spindle and slow wave amplitudes (spindles: BF_10_ = 0.37, F_(1,34)_ = 1.08, p = 0.31, ges < 0.01; slow waves: BF_10_ = 0.25, F_(1,31)_ = 0.05, p = 0.84, ges < 0.01), nor were there differences between groups (spindles: BF_10_ = 0.83, F_(1,34)_ = 0.07, p = 0.80, ges < 0.01; slow waves: BF_10_ = 0.54, F_(1,31)_ = 0.04, p = 0.84, ges < 0.01) or interaction effects (spindles: BF_10_ = 0.62, F_(1,34)_ = 1.59, p = 0.22, ges < 0.01; slow waves: BF_10_ = 0.41, F_(1,31)_ = 0.64, p = 0.43, ges < 0.01; see Figure 4.C-D).

Findings on spindle frequency are presented in **Supplementary Material, The Modulatory Effects of TMR**. In brief, no differences in these characteristics were identified during TMR and silent periods, neither in PD nor in HOA.

### Associations between Spindle and Slow Wave Characteristics and Performance Changes

The relationship between the electrophysiological metrics during NREM2-NREM3 sleep and behavioural performance was analysed using linear mixed models, including group (PD/HOA), the electrophysiological metric during the TMR periods, and the interaction between these factors as predictors of the offline changes of the reactivated sequence and of the non-reactivated sequence, separately. The results of the non-reactivated sequence and findings of the models including spindle frequency and phase-amplitude coupling as predictors are presented in detail in the **Supplementary Material, Association between Spindle and Slow Wave Characteristics and Behaviour**.

#### Spindle and Slow Wave Density

The findings show that spindle and slow wave density during the TMR periods are not associated to post-nap offline changes measured with RT and accuracy, neither of the reactivated nor of the non-reactivated sequence. Below are presented the details of the statistical tests.

##### Reaction time

Offline changes of the reactivated sequence measured with RT were not predicted by spindle density (BF_10_ = 0.43, F_(1)_ = 1.30, p = 0.26, ges = 0.03), group (BF_10_ = 0.33, F_(1)_ = 0.38, p = 0.54, ges = 0.01), nor their interaction (BF_10_ = 0.40, F_(1)_ = 0.63, p = 0.43, ges = 0.02) (see Figure 5.A, Reactivated). The analysis of the non-reactivated sequence also showed weak evidence for no main effects or interaction (group: BF_10_ = 0.33; spindle density: BF_10_ = 0.34; group by spindle density: BF_10_ = 0.47, see Figure 5.A, Non-Reactivated).

**Figure 5.**
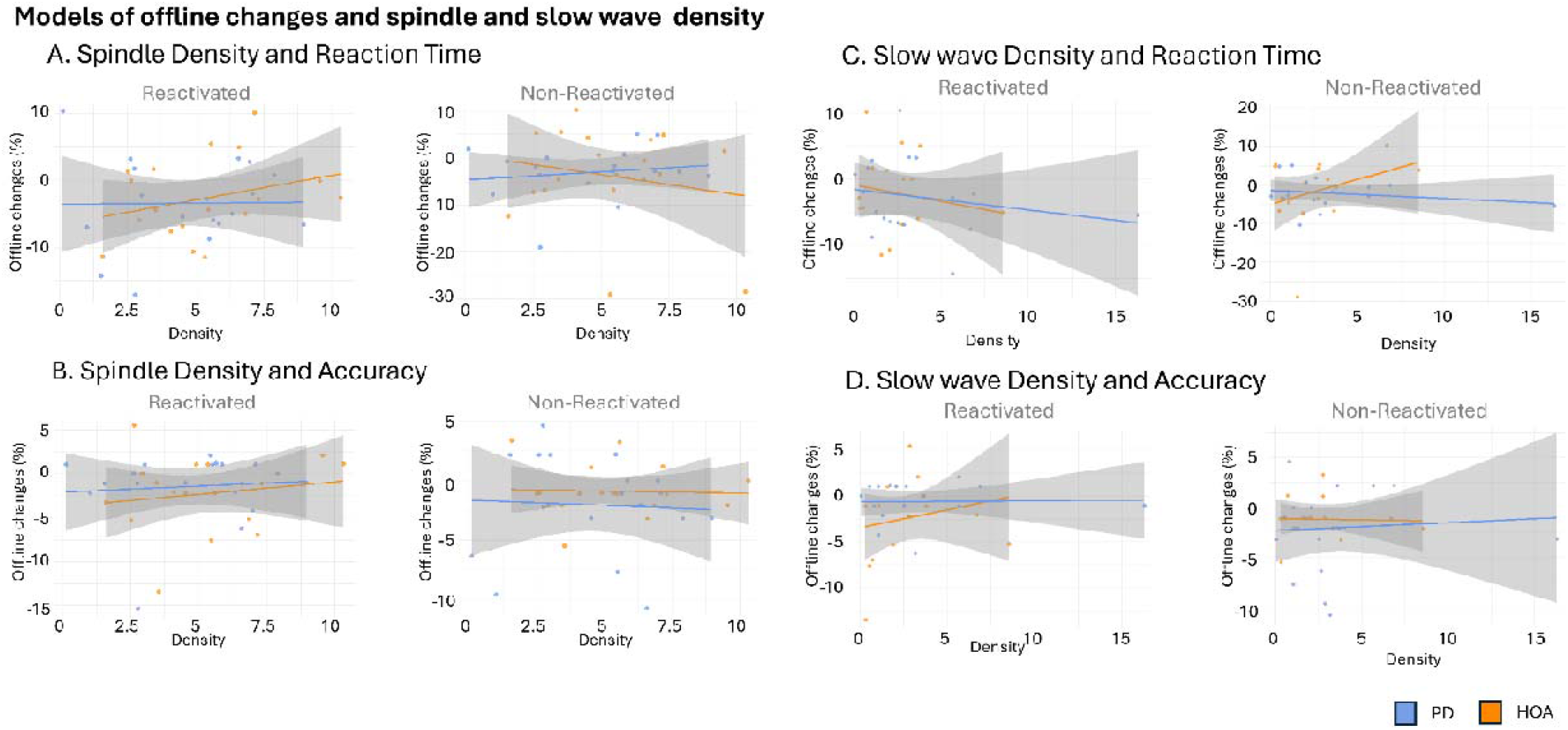
Linear mixed models for the prediction of offline relative changes post-intervention with sleep micro-architecture metrics in PD (blue) and HOA (orange). (A) Spindle density and offline changes measured with reaction time, (B) and accuracy, (C) slow wave density and offline changes measured with reaction time (D) and accuracy. Dots represent individual values, with linear regression lines for PD (blue) and HOA (orange). The grey areas indicate the standard error. Spindle density data: 17 people with PD and 17 HOA in the TMR periods, and for 17 people with PD and 17 HOA in the silence periods. Slow wave density data: 16 people with PD and 17 HOA in the TMR periods, and for 17 people with PD and 17 HOA in the silence periods.

We found weak evidence for no effect of slow wave density (BF_10_ = 0.53, F_(1)_ = 0.53, p = 0.47, ges = 0.02), group (BF_10_ = 0.34, F_(1)_ = 0.05, p = 0.83, ges < 0.01), and their interaction (BF_10_ = 0.35, F_(1)_ = 0.05, p = 0.82, ges < 0.01) on offline changes of the reactivated sequence post-nap (see Figure 5.C, Reactivated). Similarly, the analysis of the non-reactivated sequence showed weak evidence for no main effect or interaction (group: BF_10_ = 0.34; slow wave density: BF_10_ = 0.35; group by slow wave density: BF_10_ = 0.61, see Figure 5.C, Non-Reactivated).

##### Accuracy

Offline changes of the reactivated sequence measured with accuracy were not predicted by spindle density during auditory stimulation periods (BF_10_ = 0.38, F_(1)_ = 0.42, p = 0.52, ges = 0.01), group (BF_10_ = 0.37, F_(1)_ = 0.25, p = 0.62, ges < 0.01), or their interaction (BF_10_ = 0.33, F_(1)_ = 0.59, p = 0.81, ges < 0.01) (see Figure 5.B, Reactivated). Weak to moderate evidence for no effect was found for the analysis on the non-reactivated sequence (group: BF_10_ = 0.49; spindle density: BF_10_ = 0.32; group by spindle density: BF_10_ = 0.32, see Figure 5.B, Non-Reactivated).

Similarly, weak evidence was found for no effect of slow wave density during auditory stimulation periods (BF_10_ = 0.40, F_(1)_ = 0.92, p = 0.34, ges = 0.02), group (BF_10_ = 0.76, F_(1)_ = 2.33, p = 0.14, ges = 0.06), and their interaction (BF_10_ = 0.36, F_(1)_ = 0.65, p = 0.43, ges = 0.02) (see Figure 5.D, Reactivated). Weak evidence for no effect was also found in the analysis of the non-reactivated sequence (group: BF_10_ = 0.40; slow wave density: BF_10_ = 0.34; group by slow wave density: BF_10_ = 0.35, see Figure 5.D, Non-Reactivated).

#### Spindle Amplitude

Overall, spindle amplitude was not associated to post-nap offline changes of the reactivated and of the non-reactivated sequence, measured both with RT and accuracy. Detailed statistical analyses are reported below.

##### Reaction time

The model highlighted weak evidence for no effect of group (BF_10_ = 0.33, F_(1)_ = 0.97, p = 0.33, ges = 0.03), spindle amplitude (BF_10_ = 0.33, F_(1)_ = 0.28, p = 0.60, ges < 0.01) and their interaction (BF_10_ = 0.51, F_(1)_ = 1.20, p = 0.28, ges = 0.03) on offline changes post-nap (see Figure 6.A, Reactivated). Weak evidence for no effect was also found for the analysis on the non-reactivated sequence (group: BF_10_ = 0.33; spindle amplitude: BF_10_ = 0.56; group by spindle amplitude: BF_10_ = 0.47, see Figure 6.A, Non-Reactivated).

**Figure 6.**
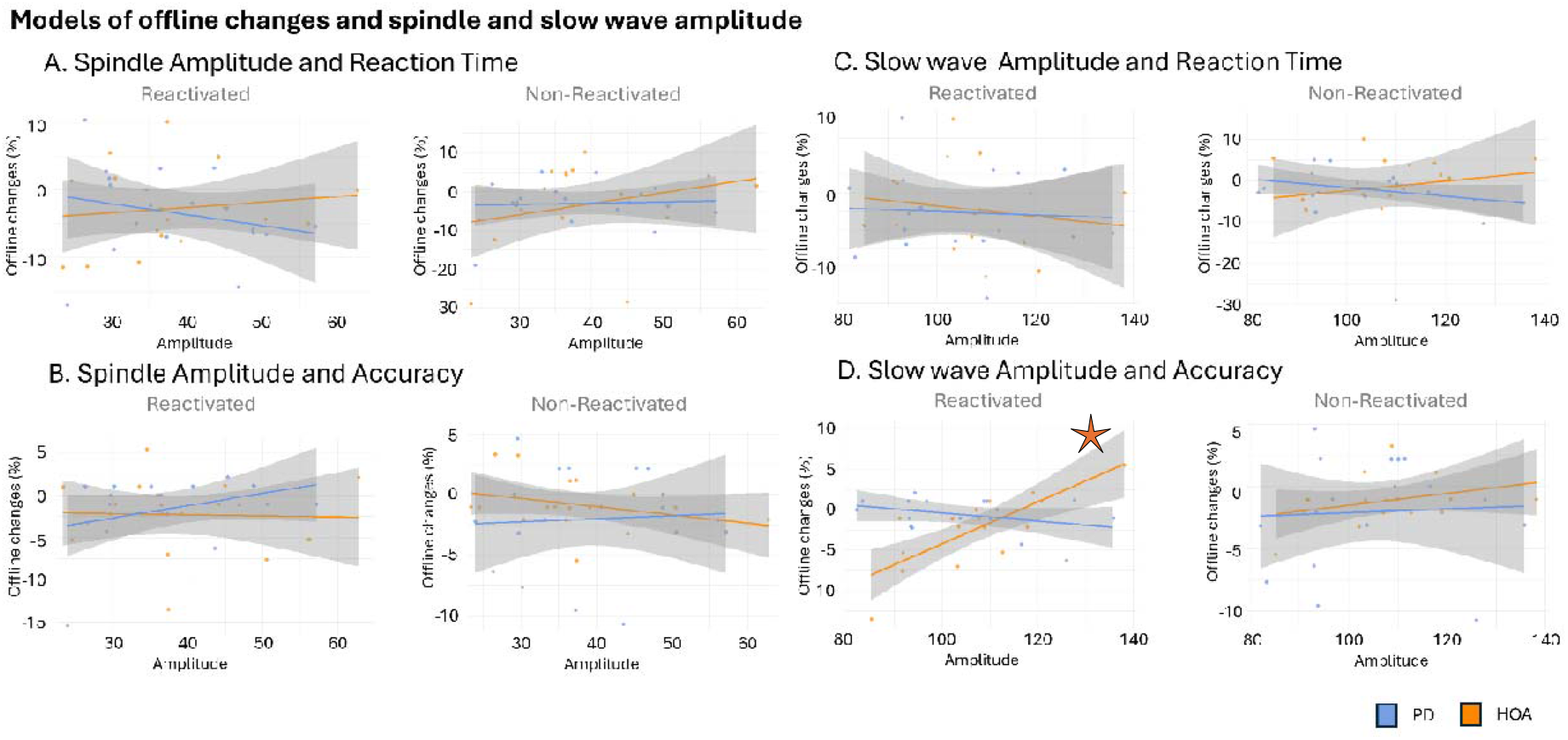
Linear mixed models for the prediction of offline relative changes post-intervention with sleep micro-architecture metrics in PD (blue) and HOA (orange). (A) Spindle amplitude and offline changes measured with reaction time, (B) and accuracy, (C) slow wave amplitude and offline changes measured with reaction time (D) and accuracy. Dots represent individual values, with linear regression lines for PD (blue) and HOA (orange). The grey areas indicate the standard error. Spindle amplitude data: 17 people with PD and 17 HOA in the TMR periods, and for 17 people with PD and 17 HOA in the silence periods. Slow wave amplitude data: 16 people with PD and 17 HOA in the TMR periods, and for 17 people with PD and 17 HOA in the silence periods. * significant predictive value of the sleep metric considered for offline changes

##### Accuracy

The main effects of group (BF_10_ = 0.37, F_(1)_ = 0.85, p = 0.36, ges = 0.02), spindle amplitude during TMR application (BF_10_ = 0.42, F_(1)_ = 0.02, p = 0.89, ges < 0.01) or their interaction (BF_10_ = 0.49, F_(1)_ = 1.21, p = 0.28, ges = 0.04) were not predictive of performance changes measured with accuracy (see Figure 6.B, Reactivated). Similar weak evidence for no effect was highlighted by the analysis on the non-reactivated sequence (group: BF_10_ = 0.49; spindle amplitude: BF_10_ = 0.35; group by spindle amplitude: BF_10_ = 0.42, see Figure 6.B, Non-Reactivated).

#### Slow Wave Amplitude

The results show that, while offline changes measured with RT did not appear to be associated to slow wave amplitude during the TMR periods, the offline changes of the reactivated sequence were positively predicted by the slow wave amplitude during TMR in HOA. This was not true for PD, nor for the offline changes of the non-reactivated sequence, measured with accuracy, in neither group. Detail of the statistical tests are reported below.

##### Reaction time

We found weak evidence for no effect of group (BF_10_ = 0.34, F_(1)_ = 0.12, p = 0.73, ges < 0.01), slow wave amplitude (BF_10_ = 0.36, F_(1)_ = 0.38, p = 0.54, ges = 0.01) and their interaction (BF_10_ = 0.34, F_(1)_ = 0.10, p = 0.75, ges < 0.01) on offline changes post-nap of the reactivated sequence (see Figure 6.C, Reactivated), and similar findings were highlighted by the analysis of the non-reactivated sequence (group: BF_10_ = 0.34; spindle amplitude: BF_10_ = 0.34; group by spindle amplitude: BF_10_ = 0.66, see Figure 6.C, Non-Reactivated).

##### Accuracy

Findings highlighted strong evidence in favour of an interaction effect of group and slow wave amplitude during TMR periods on accuracy offline changes of the reactivated sequence (BF_10_ = 29.24, F_(1)_ = 22.87, p < 0.01, ges = 0.17). Additionally, the model revealed weak evidence for no effect of slow wave amplitude (BF_10_ = 0.96, F_(1)_ = 27.36, p < 0.01, ges = 0.20) and weak evidence in favour of no effect of group (BF_10_ = 0.76, F_(1)_ = 25.28, p < 0.01, ges = 0.19). Post-hoc analyses revealed a statistically significant difference between groups (estimate of the difference between slopes = 0.31, p < 0.01), with a positive significant association between slow wave amplitude and performance changes in HOA (β = 0.26, 95% CI [0.16 – 0.36]), while this was negative and non-significant in PD (β = -0.05, 95% CI [-0.13 – 0.03]), see Figure 6.D (Reactivated). In contrast, the same analysis on the non-reactivated sequence revealed weak evidence for no main effect or interaction (group: BF_10_ = 0.40; slow wave amplitude: BF_10_ = 0.41; group by slow wave amplitude: BF_10_ = 0.34), see Figure 6.D (Non-Reactivated).

#### Phase-amplitude Coupling and Associations with Behaviour

The differences between the coupling of slow waves (0.3 – 1.5Hz frequency band) and spindles (10 – 16Hz) during TMR periods and silence, in PD and HOA were studied with Watson’s U^2^ test for homogeneity of two samples of circular data. The findings highlighted no significant difference between PD and HOA in the phase-amplitude coupling, neither during the auditory stimulation periods (U^2^= 0.15, critical U^2^ = 0.19, p > 0.10, Figure 7, TMR), nor during the silence periods (U^2^= 0.10, critical U^2^ = 0.19, p > 0.10, Figure 7, no-TMR). Within-group comparisons between auditory stimulation periods and silence periods did not show significant differences (PD: U^2^= 0.07, critical U^2^ = 0.19, p > 0.10; HOA: U^2^= 0.11, critical U^2^ = 0.19, p > 0.10).

**Figure 7.**
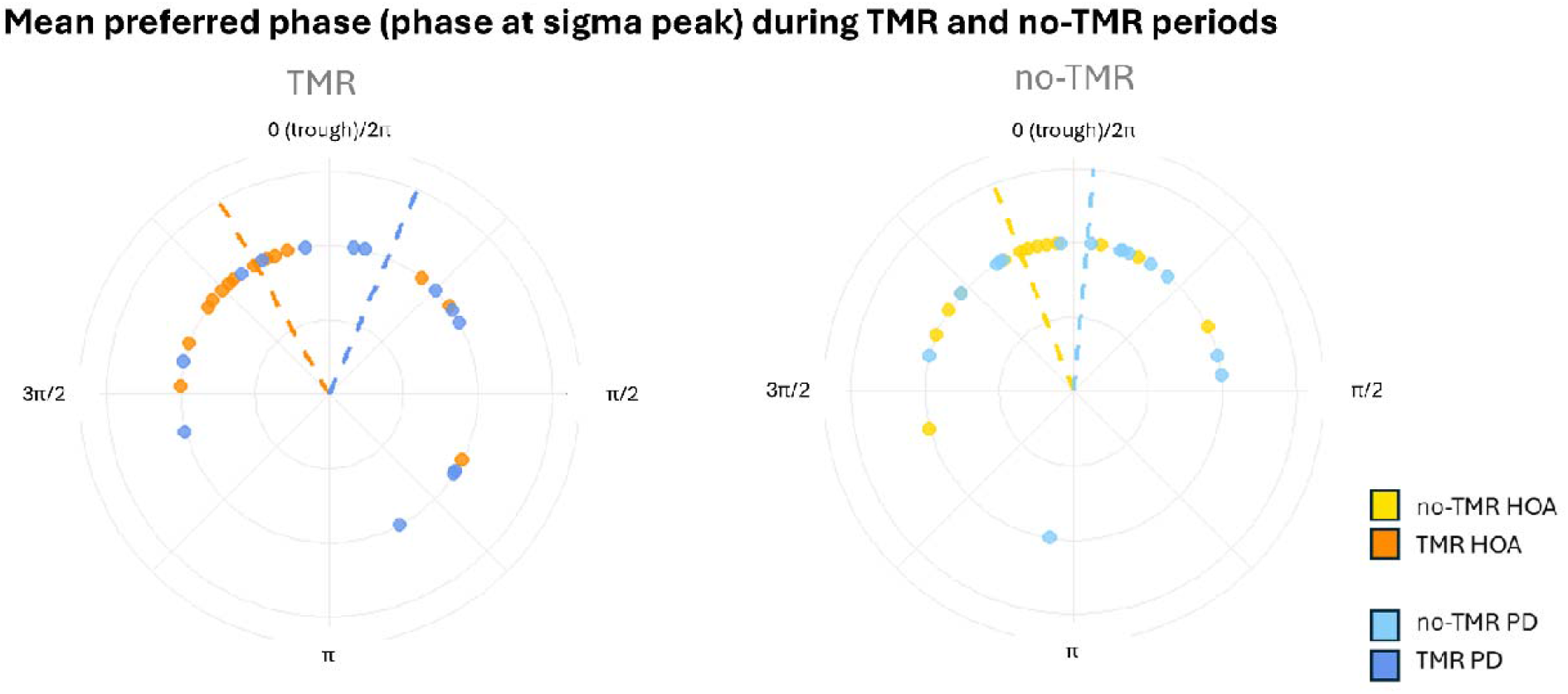
Phase at sigma peak of the mean preferred phase, computed in C3, during TMR periods and during silence periods (no-TMR). Data is presented in radians, for 13 people with PD and 14 HOA in the TMR periods, and for 13 HOA and 15 people with PD in the silence periods. Dots represent single values of mean per subject, dashed lines indicate the overall group mean.

Frequentist correlational analysis between offline changes post-nap, measured as RT, and phase at sigma peak during the auditory stimulation periods were non-significant after FDR-correction (PD reactivated: R^2^ = 0.21, p = 0.12, FDR-corrected p = 0.37; HOA reactivated: R^2^ = 0.08, p = 0.40, FDR-corrected p = 0.46; PD non-reactivated: R^2^ = 0.13, p = 0.27, FDR-corrected p = 0.39; HOA non-reactivated: R^2^ = 0.23, p = 0.07, FDR-corrected p = 0.37). Also the correlational analyses between the offline changes measured with accuracy and phase at sigma peak did not show significant differences (PD reactivated: R^2^ = 0.17, p = 0.19, FDR-corrected p = 0.37; HOA reactivated: R^2^ = 0.04, p = 0.63, FDR-corrected p = 0.63; PD non-reactivated: R^2^ = 0.18, p = 0.16, FDR-corrected p = 0.37; HOA non-reactivated: R^2^ = 0.11, p = 0.29, FDR-corrected p = 0.39).

Overall, the phase at sigma peak was not different between people with PD and HOA, nor between auditory stimulation and silence periods within-groups, and no evidence for an association between the phase at sigma peak and post-nap offline performance changes was found, neither for the reactivated sequence, nor for the non-reactivated sequence.

## Discussion

In this study, we investigated how performance on a motor sequence learning task is modulated by targeted memory reactivation provided during NREM2 and NREM3 sleep of a 2-hour post-learning nap in people with PD and HOA. We also investigated the sleep-related electrophysiological changes induced by the auditory stimulation presented during the nap, and their association with performance changes. The application of TMR did not yield an improvement of motor performance of the reactivated sequence, compared to the non-reactivated one, neither immediately after the 2-hour nap nor after 24 hours retention. However, the application of the auditory stimuli did modulate both spindle and slow wave densities. Specifically, slow wave density increased while spindle density decreased during the periods of sleep with auditory cues in both groups. Additionally, slow wave amplitude was positively associated to performance improvements of the reactivated sequence in HOA, more than in PD, where such relationship was not observed. No such effect was observed for the non-reactivated sequence.

People with PD and HOA showed similar post-nap performance changes, with no differences between the reactivated and the non-reactivated sequence. These findings are in agreement with a previous study on the use of TMR on memory consolidation of a serial reaction time task (SRTT) in HOA (Nicolas et al., 2024), suggesting a possible non-specific effect of TMR. Here, we further show that a similar effect is observable in PD. Such similarity in post-nap performance changes between PD and HOA is also in line with previous studies investigating the immediate effects of sleep on motor sequence learning in PD and HOA (Lanir-Azaria et al., 2024; Micca et al., 2025; Terpening et al., 2013). Conversely, the lack of performance differences at 24-hour retention between the groups is in disagreement with the work of Terpening et al. showing a failure of people with PD to show long-term retention after a night of sleep post-practice (Terpening et al., 2013). However, such difference between groups was not evidenced in a recent study from our team (Micca et al., 2025), where performance at 24-hour retention was similar between groups. These contrasts across studies could be explained by the amount of practice provided, which has been identified as a contributing factor to later retention both in PD and HOA (Cristini et al., 2023; King et al., 2016). Specifically, in the study by Terpening et al., initial learning consisted of twelve 30-second practice blocks of finger tapping task, and retest comprised six 30-second blocks. The current study implemented 12 to 14 practice and test blocks for each sequence at each session, accounting for both single and dual-task blocks. When additional practice is implemented, HOA can present greater sleep-related performance gains (King et al., 2016), while people with PD may be limited due to the cumulative retention deficits. It appears that the effect of sleep alone on retention is weaker in the ageing population than in healthy young adults, and it could be shadowed by the benefit of practice. In PD, immediate benefits from added practice may be similar to HOA, as well as sleep-related consolidation processes, but the striatal dysfunction may hinder the long-term benefits and possible automaticity of the skill learnt. Hence, while sleep interventions alone may not be sufficient to aid retention in PD, their combination with added task practice could contribute to improvements in retention similar to HOA.

Moreover, PD patients seem to show task-specific deficits in motor memory consolidation, with larger consolidation impairments evidenced with sensorimotor and visuospatial tasks, as compared to motor sequence learning tasks (Cristini et al., 2023). This could be explained by the fact that MSL tasks largely involve cortical regions and thalamus, in contrast with sensorimotor tasks, which show greater involvement of the basal ganglia (Hardwick et al., 2013) that are impaired in PD. This interpretation is further supported by results reported by Van Roy et al. (Van Roy et al., 2025), where the interaction between practice and consolidation at the micro-scale level was studied. Their findings show that online gains are significantly lower while offline gains are larger than HOA suggesting striatal-cortical deficits involved in practice subsequently compensated by hippocampal-cortical activity during offline periods. These speculations could explain the lack of differences between people in the early stages of PD and HOA, where not only cortical, but also hippocampal involvement would hold a compensatory role in the sequence representation, and this might be further enhanced by sleep. Importantly, the cortical involvement is particularly evidenced when the sequence is explicit, and although a SRTT was used in the current study, all participants were priorly informed that two distinct sequences were presented. Hence, people with PD could have activated compensatory executive circuits that bypass the affected striatal-cortical circuits involved in implicit motor performance, thus further minimising differences with HOA post-nap and at 24-hour retention.

Importantly, we observed a slower learning rate in PD compared to HOA at initial learning. However, the improvement over time was comparable between groups at post-nap and after 24 hours. This finding is in disagreement with our previous research on the effects of a 2-hour nap on consolidation, where PD and HOA showed similar online learning rates pre-nap and post-nap, but not at 24-hour retention, where HOA showed overall better performance compared to PD (Micca et al., 2025). Such discrepancies might be explained by the requirement to learn two distinct sequences in the current study, which could have affected the strength of memory trace encoding prior to the intervention.

The nature of the task used could also explain the lack of differences in dual-task costs between populations. These findings are in line with our previous work showing that performance of an MSL task was similarly affected by a working memory task both in people with PD and HOA (Micca et al., 2025). Yet, it has been demonstrated that performance of a writing task was affected by a working memory task in people with PD, compared to HOA (Broeder et al., 2014). Previous research suggested that people with PD can show pseudo-automaticity in the early stages of the disease, as cortical regions may compensate for the dysfunctional striatum when having to maintain performance with an additional distraction task (Nackaerts et al., 2019; Nieuwhof et al., 2017). Our findings suggest that the SRTT may not be cognitively challenging enough to elicit dual-tasking impairments often observed in PD (Broeder et al., 2014; Cristini et al., 2023; Ozcan Gulsen & Tunca Yilmaz, 2023; Proud & Morris, 2010). It is also possible that disease-related factors in people with PD limited the group differences, as compensatory mechanisms could have allowed them to perform the sequences with minimal dual-task interference. In other words, the secondary counting task applied might have not been effectively challenging to induce attentional interference.

Remarkably, auditory stimulation during sleep did induce modulation of spindles and slow wave characteristics, which have been widely associated to sleep-related consolidation processes, similarly in both groups. Specifically, the increase of slow wave density during the TMR periods has been consistently demonstrated, with auditory stimuli having a slow-wave entraining effect in healthy young (Wunderlin et al., 2021), older adults (Lustenberger et al., 2022; Nicolas et al., 2024), as well as in PD, as recently demonstrated by Schreiner et al. (Schreiner et al., 2026) during overnight sleep. Strikingly, This effect was particularly evident when stimulation was applied during the first 2-3 hours of the night (Schreiner et al., 2026), a time similar to the 2-hour napping opportunity offered in the current study. Conversely, spindle density was reduced during the TMR periods, when compared to the silent periods, both in PD and in HOA. These similarities between populations are in line with our previous research (Micca et al., 2025) showing comparable sleep micro-architecture characteristics between PD and HOA during a 2-hour post-learning nap. Moreover, these effects of TMR have been also demonstrated in healthy young (Nicolas et al., 2022) and older adults by (Nicolas et al., 2024), where spindle density was decreased upon presentation of an auditory stimulus, while the slow-wave spindle synchronised coupling was enhanced. Our findings demonstrate that replaying sounds during sleep induces spindle and slow-wave modulation in PD, suggesting that patients with HY 2-2.5 have underlying electrophysiological mechanisms during NREM2-NREM3 sleep that are comparable to HOA.

By contrast, no differences were found between TMR periods and silent periods on spindle and slow wave amplitude, spindle frequency or phase-amplitude coupling in neither group. Specifically, a peak in the spindle amplitude close to the trough of the slow wave was found, which was not significantly modulated by the TMR cues. It has long been postulated that neuroplasticity peaks during the depolarisation up-state of the slow wave (Born & Wilhelm, 2012), and a recent meta-analysis showed that the phase-amplitude coupling has a central role in memory consolidation (Ng et al., 2025). The same meta-analysis also showed that ageing is a contributing factor to the uncoupling between spindles and slow waves, with consequences for memory consolidation (Helfrich et al., 2018; Ng et al., 2025). Specifically, ageing was characterized by an anticipation of the sigma peak relative to the slow wave peak, in contrast to young adults, in whom it occurs in close temporal proximity to or shortly after the slow wave peak (Helfrich et al., 2018). The PAC during TMR and silent periods was not different in the present study, nor did we find differences between groups, suggesting that TMR alone may not contribute to sigma peak amplitude shift towards the slow wave up-state. Moreover, PAC during TMR periods was not associated to offline performance changes neither of the reactivated nor the non-reactivated sequence, both in PD and in HOA, further suggesting that age-related consolidation impairments may hinder memory consolidation processes during sleep rather than PD-specific degeneration.

Yet, a differential association between slow-wave amplitude during TMR periods and accuracy improvements was found between populations, where in HOA, but not in PD, slow wave amplitude was positively associated to accuracy of the reactivated sequence only. This result is partially in agreement with the work of (Terpening et al., 2013), where NREM3 sleep stage duration predicted performance improvements in HOA, but not in people with PD. Since slow-wave amplitude has been previously associated to consolidation in young adults (Heib et al., 2013), it is possible that TMR in HOA may promote synchronisation in the task-related cortical areas, with underlying consolidation mechanisms similar to healthy young adults (Nicolas et al., 2022, 2024). Nonetheless, associations of TMR-related slow-wave modulation and offline changes were not observed in PD. This result is striking, as in our earlier work investigating the effect of a nap on motor memory consolidation, we did find a positive correlation between slow wave amplitude and offline changes in PD, although not significantly different from HOA (Micca et al., 2025).

While spindle and slow wave density differed between TMR and silent periods, these were not associated to better offline changes. This is in line with the findings by (Nicolas et al., 2024), suggesting that these metrics may have a non-specific effect on the reactivation of the newly acquired memory traces. In a similar way, we did not find any additional modulatory effect of TMR on spindle amplitude and frequency, nor an association of these metrics with consolidation. These results are in agreement with the study by Fogel et al. (2017) (S. Fogel et al., 2017), indicating that alterations in spindle characteristics can be associated to age-related consolidation deficits.

Importantly, this study presents some limitations. Firstly, we did not investigate the possible effect of TMR during sleep compared to a wake control group, therefore possible sleep-related and wake-related memory consolidation processes were not compared. It is noteworthy that in the current study people with PD were always tested in their usual ON-medication state, a factor that could have masked PD-specific deficits (Ruitenberg et al., 2015). Further, to reduce circadian interference and for feasibility reasons, a nap paradigm was used. While TMR during 2-hour afternoon sleep opportunity could enable consolidation effects during NREM2 and NREM3 sleep, its effects during a full night of sleep, and in the OFF-medication state, which people with PD can experience overnight, requires further investigation. Moreover, the melodies paired to the two sequences contained the same tones in a different order, and may have been too similar to elicit optimal association to the individual motor memory trace. Additionally, the lack of a control sound could not elucidate whether the modulatory effects here found were related to TMR or to mere auditory stimulation. In the future, the use of singular sounds on different octaves, and the integration of a control sound, non-associated to previously learnt sequences, could facilitate the differentiation between stimuli, and may elicit clearer differences between reactivated and non-reactivated sequence. Finally, the task used in this study could have prevented from observing performance differences between PD and HOA. Further research is thus required to test the effectiveness of TMR on sensorimotor or visual adaptation task where patients can show larger deficits may provide clearer answers on the PD-specific differences in sleep-dependent memory consolidation processes.

On the whole, these findings suggest that auditory stimulation with a sound that was previously associated to a memory trace has a modulatory effect on spindles and slow waves both in PD and HOA, similar to what has been shown in healthy young adults. They also suggest that SW amplitude is associated to increase in performance in HOA and that auditory stimulation auditory stimulation can enhance SW amplitude.. Future work is needed to examine the effects of TMR after more extensive initial learning and across other tasks. In addition, further efforts to optimize TMR protocols and to enhance their impact on consolidation through the modulation of sleep oscillations will be essential to establish TMR as a beneficial memory-protective adjunct to Parkinson’s rehabilitation.

## Materials and Methods

### Study Design

The protocol for this phase-II within-subject randomised controlled trial was approved by the Medical Ethical Committee Research UZ/KU Leuven (S61792). Experimental procedures were conducted in accordance with the declaration of Helsinki and the World Medical Association, 2013 (‘World Medical Association Declaration of Helsinki: Ethical Principles for Medical Research Involving Human Subjects’, 2013) and all participants provided written informed consent prior to the study. The procedures and main hypotheses were pre-registered on clinicaltrials.gov (Experiment 2, clinicaltrials.gov/study/NCT04144283), and the reporting is in agreement with the Consolidated Standards of Reporting Trials (CONSORT) guidelines (see **Supplementary Material, CONSORT 2010 checklist of information to include when reporting a randomised trial**) (Schulz et al., 2010). We report the deviations from preregistration in **Supplementary Material, List of Deviations from Pre-registration**. The screening took place at the Centre for Sleep Monitoring at the University Hospital Leuven and the TMR experiment took place at the Brain Dynamics Laboratory (Brainshub) of KU Leuven in Belgium.

The experiment is part of a larger project comprising another study on the effects of napping compared to wakefulness on motor memory consolidation in PD (Micca et al., 2025). Following a screening night and nap experiment as part of another study (Micca et al., 2025), 40 participants (20 people with PD and 20 HOA) expressing interest in the current study were consecutively recruited for this second experiment. We accounted for a minimum of 4 weeks between the previous and current experiment to minimise carry-over effects. However the time in-between was much longer for several participants due to COVID19 (>10 months for 26 participants, with a maximum of 31 months). Inclusion occurred consecutively and the data collection was considered completed after the final preregistered sample of N =20 per group was achieved. No adverse events were reported.

Inclusion criteria for all participants were: (1) right-handedness, assessed with the Edinburgh Handedness Inventory (with laterality quotient > 40)(Oldfield, 1971); (2) ≥40 years of age (Quinn et al., 1987); (3) Mini Mental State Examination (MMSE) score ≥24 (Tombaugh & McIntyre, 1992). Exclusion criteria for all participants were: (1) known diagnosis of insomnia, to maximize the chance of sleep during the experimental nap with TMR; (2) severe apnoea (Apnoea Hypopnea Index (AHI) ≥30, accounting for obstructive, central and mixed apnea together) as determined by an overnight screening polysomnography (PSG), to minimise sleep disruption while still allowing for broad inclusion of people with PD and HOA, who can often present sleep disordered breathing (Aini et al., 2025; Maggi et al., 2024); (3) presence of other known disease or condition that could significantly interfere with the experimental procedures. PD-specific additional inclusion criteria were: (1) a diagnosis of PD made by a neurologist, according to the latest Movement Disorder Society diagnostic criteria (Postuma et al., 2015) and (2) Hoehn & Yahr stage (H&Y) I-III ON medication (Hoehn & Yahr, 1967). The PD-specific exclusion criteria comprised: (1) deep brain stimulation, since its influence on memory consolidation processes is still unclear; (2) self-reported freezing of gait (FOG) with more than one freezing episode per month, according to the New Freezing of Gait Questionnaire (NFOGQ) (Nieuwboer et al., 2009), given that people with PD and freezing of gait have more prominent motor learning issues, and to minimise the possibility of freezing of the upper limb during performance of the motor task (Vercruysse et al., 2014).

### General Experimental Protocol

The present study utilised a between- and within-subject design. Participants were consecutively recruited for this experiment, consisting of three test sessions across two days (see Figure 1). Before the screening night, general demographic characteristics were collected and the MMSE and NFOGQ were administered. Data of sleep disordered breathing were then gathered with the overnight PSG, allowing to calculate the apnoea hypopnea index (AHI). Given that the study was disrupted by the COVID-19 pandemic, the test sessions were performed at the earliest 3 months and at the latest 35 months after the screening night. The screening nights were not repeated due to lack of resources during COVID19, but sleep was monitored for the 7 days before the experiment with a sleep diary (data not reported) and the MDS-UPDRS and the MoCA were re-administered.

At the beginning of each test session, participants performed a Psychomotor Vigilance Test (PVT) (details presented in **Supplementary Material, Psychomotor Vigilance Test**) to assess their objective level of vigilance (Dinges & Powell, 1985). Prior to initial encoding on the first session (pre-intervention, ∼10:30 a.m. of experiment Day 1), participants were tested on a random bimanual serial reaction time task (SRTT) to assess baseline motor execution. This was followed by the training and testing of 2 sequential bimanual SRTTs of eight elements each (see section below for motor task details). Each visual stimulus of the SRTT was associated to a distinct sound, therefore the two sequences, when performed, generated two distinct melodies on the same octave. After the pre-intervention session, participants ate their packed lunch, and they were subsequently offered a 2-hour opportunity to sleep. During the stages NREM2 and NREM3 of the 2-hour nap opportunity, TMR was applied by replaying one of the two melodies associated to one of the two previously practiced motor sequences (detailed below in Motor Tasks). During the nap, all participants were monitored with low-density PSG (detailed below in Polysomnography and Targeted Memory Reactivation). The second session (post-nap) started 30-40 minutes after the 2-hour nap opportunity period, to allow for possible sleep inertia effects to dissipate (Tassi & Muzet, 2000). During the post-nap session, the extended training and testing of the same sequences were again performed, and served as immediate post-nap assessment. These were then followed by a dual-task test, consisting of the performance of the same sequential SRTTs, with an additional counting task, and a random SRTT testing, which was performed at the end of the post-nap session. Lastly, a third session (post-night) took place on Day 2, 24 hours after the post-nap session, with experimental procedures identical to the post-nap session. Between Day 1 and Day 2, participants slept at home, monitored with a sleep diary (data presented in Table 1 and Actiwatch (data not reported due to a technical issue during the processing).

Randomisation of the first sequence to-be-trained and of the sequence to-be-reactivated during the nap with TMR were performed 1:1 for PD and HOA separately with block size 4 and was stratified for age (under 65 years vs. 65 years or older) and self-reported biological gender (male vs. female). These randomisations were performed using a computerised blocked randomisation sequence (app.studyrandomizer.com). The researcher shared the concealed assignments with the study assessors via secured email.

Finally, participants underwent the Montreal Cognitive Assessment (MoCA) and the Movement Disorder Society Unified Parkinson Disease Rating Scale (MDS-UPDRS) at the beginning of the third experimental session (Day 2).

### Motor Tasks

All the tasks were scripted in Matlab (Math Works Inc, Natick, MA, USA; version R2018a), using the Psychophysics Toolbox extensions (Brainard & Vision, 1997; Kleiner et al., 2007; Pelli & Vision, 1997). Participants were sitting comfortably in front of a laptop screen and used a keyboard (model: Accuratus, Monster 2, QWERTY) to practice the SRTTs. The keyboard was adapted by removing all the keys except for the A, S, D and F keys, used with the left non-dominant hand during the task, and the J, K, L, and “;” keys, used with the right dominant hand, and the spacebar, used during the Psychomotor Vigilance Test (PVT).

Both the random and the sequential SRTTs consisted of visually cued bimanual finger-tapping tasks. For the sequential SRTT, two sequences were practiced, alternating between blocks of sequence A (4-7-3-8-6-2-5-1, with 1 corresponding to the pinky finger of the left hand, and 8 corresponding to the pinky finger of the right hand, in a left-to-right progression, without the thumbs) and sequence B (7-2-8-4-1-6-3-5). The visual cues consisted of 8 squares, represented in black with green contour during the practice blocks, one for each corresponding finger positioned on the keyboard, and they were displayed horizontally in the centre of the laptop screen, on a black background. During the practice blocks, the area of the squares would light up sequentially in green when the corresponding key of the sequence was to be pressed. The following visual cue of the sequence, paired with the corresponding auditory tone, would then light up/be played only after the key was pressed in response of the previous cue with no interstimulus interval, irrespective of the correctness of the previous key press. The squares were represented in black with red contour during the break periods of the duration of 20s between practice blocks. For an additional period of 3s before the start of each practice block, all the squares were represented in black with green contour, and this period served as a prompt for the participants to prepare to perform the pending task. A specific auditory tone was presented simultaneously to the visual cues during practice, so that each bimanual sequential SRTT was associated to a distinct melody of 8 sounds.

The sequential SRTT training consisted of 20 blocks of 48 key presses (i.e., ideally corresponding to 6 sequences per block), whereby the two sequences were alternated between blocks, for a total of 10 training blocks per sequence. After training, all participants had a 2-minute break to allow fatigue dissipation (Pan & Rickard, 2015), followed by another four practice blocks (i.e., ‘test’), again alternating sequence A and B between blocks, for a total of two test blocks per sequence.

The random SRTT, also cued with sounds corresponding to the visual cues appearing on the screen, served as a control condition to determine if any improvement observed on the sequential SRTT was related to actual sequence learning or to general improvement in motor execution. The random SRTT was composed of 4 blocks of 48 consecutive key presses each and was performed before the sequential SRTT of the pre-nap session and after the sequential SRTT of the post-nap and post-night sessions.

During both the random and the sequential SRTTs, participants were instructed to press keys as quickly and as accurately as possible in response to the visual and auditory cues. No feedback on performance was provided during or after the training and testing. During these tasks, the reaction time (RT) of each key press and the accuracy of the response were recorded for data processing.

The dual-task version of the sequential SRTT task was performed after the test of the single-task SRTT, at post-nap and post-night. For this task, participants had to mentally keep count of how many times the squares lit up in white rather than in green (occurring at 5 to 8 pseudo-random moments during each block of 48 keys), while continuing to perform the task. At the end of each block, they verbally reported the count to the assessor to verify that they were actively engaged in the secondary task. The verbal responses were logged, but not further analysed.

### Auditory Stimulation

Each visual cue was associated to an auditory note (C4-D4-E4-F4-G4-A4-B4-C5, where the letter indicates the note, and the number indicates the octave). Each of these notes was paired with an ordinal position of the elements of the sequence (i.e., key 1 – C4; key 2 – D4, etc.). Consequently, the two motor sequences corresponded to two distinct melodies. One of these melodies was subsequently replayed when NREM2 and NREM3 sleep stages were detected during the 2-hour nap opportunity. Sleep stage detection was performed online by an experienced researcher. The sounds were stopped whenever N1, REM or wake were detected. These auditory stimuli were presented via noise cancelling earplugs (Etymotic Research Inc. – 3C), both during task practice and during the nap.

Prior to task practice, on the first session, all participants underwent a minimal auditory threshold test, where the volume of each auditory tone was established with a transformed one-down one-up procedure (Leek, 2001; Levitt, 1971). The volume set during task practice was set at 1000% pressure of the determined minimal auditory threshold, while the pressure was set to 140% when the auditory stimuli were replayed during the nap, to ensure that participants would not be awoken (Nicolas et al., 2024; Sterpenich et al., 2014).

### Polysomnography and Targeted Memory Reactivation

Monitoring of the experimental naps was performed with a digital sleep recorder (V-Amp, Brain Products, Gilching, Germany). The electroencephalographic (EEG) setup comprised recordings made from electrodes Fz, Cz, Pz, F3, F4, C3, C4, O1, O2, according to the international 10–20 system. Fpz constituted the ground and A2 was used as recording reference. Electro-oculographic (EOG) recordings were made with two electrodes placed 1 cm from the external-superior canthus of the right eye, and external-inferior canthus of the left eye, while electromyography was recorded with three electrodes, one placed on the chin, and two placed respectively on the right and left side of the submandibular muscle. Impedances for the electrodes were kept below 10 kΩ and data of EEG, EOG and EMG were collected with a sampling frequency of 1000 Hz.

During the experimental nap, visual online detection of NREM2-NREM3 sleep stages was performed by an experienced researcher, following the guidelines from the American Academy of Sleep Medicine (Berry et al., 2018). Upon online detection of NREM2-NREM3 stages, auditory sequences were presented 8 consecutive times, with a silent period of 2s between each sequence presentation. A rest period of 1 minute followed each group of 8 auditory sequences presentations. The interstimulus interval was individualised, and it was determined as the average RT of the 4 blocks of the sequential SRTT (test) preceding the experimental nap (Veldman et al., 2021). The TMR and EEG synchronisation failed in four participants (two HOA and two PD). Although TMR was applied, we could not distinguish periods of sequence replay and periods of silence in these participants. Their data were thus excluded for the EEG analyses, but the behavioural data was analysed as per an *intention-to-treat*.

### Data Analysis

Statistical tests were performed with R open-source software (R Core Team, 2022). Given the modest sample size of this phase-two trial, we implemented Bayes factors in the reporting of the findings, to give an overview of the likelihood that the observed data supported the acceptance of the alternative hypothesis (e.g., the two conditions have different means/variances) or the null hypothesis (e.g., the two conditions have similar means/variances). Bayes factors (BF) were computed using the BayesFactor package (Morey et al., 2022), and these were interpreted according to the guidelines provided by Lee and Wagenmakers (Lee & Wagenmakers, 2014) with BF>1 indicating evidence in favour of the alternative hypothesis and BF<1 suggesting evidence in favour of the null hypothesis. Post-hoc comparisons were performed when BFs suggested at least moderate evidence in favour of the rejection of the null hypothesis (i.e., BF > 3). For the analyses using Bayesian statistics, (i) a noninformative Jeffreys prior for the variance and a Cauchy prior for the standardised effect size with value √2/2 were assigned, (ii) 10.000 Markov Chain Monte Carlo iterations were used, and (iii) missing data were excluded (Morey et al., 2022) (missing data, if present, are reported for each specific computed metric).

Bayesian analyses of variance (ANOVA), Bayesian t-tests and Bayesian linear mixed models were performed with the package BayesFactor (Morey et al., 2022). The findings of these analyses were complemented with the results of frequentist statistics. The equivalent tests with frequentist ANOVAs were implemented in the ez package in R (Lawrence, 2016), where Greenhouse-Geisser (GG) correction was applied in case of violation of the sphericity assumption. Bayesian t-tests were complemented with Student’s t-tests or Mann-Whitney U tests. Frequentist linear mixed models complemented their Bayesian counterpart. Of these frequentist tests we report the test results (Fisher’s F, Student’s t, Wilcoxon W) confidence intervals when applicable, p-values, and effect sizes, expressed as generalised eta squared (ges) (Bakeman, 2005), Cohen’s d (for frequentist parametric tests) or rank-biserial correlation (r_rb_) for frequentist non-parametric tests.

### Behaviour

#### Processing

The primary metric used for the calculation of performance changes was reaction time (RT). This was measured as the time in seconds between cue appearance on the screen and the subsequent correct keypress.

For the statistical analyses, outliers were identified using the Tukey method (Tukey, 1960) that leverages interquartile ranges, given the non-normal distribution of the data from RT. Values of RT falling above or below 1.5 interquartile range were thus considered outliers (PD random sequence: 2.50%; PD training reactivated: 0.87% - non-reactivated: 0.95%; PD test reactivated: 0.17% - non-reactivated: 0.18%; PD dual-task reactivated: 0.14% - non-reactivated: 0.14%; HOA random sequence 2.37%; HOA training reactivated: 0.83% - non-reactivated: 0.78%; HOA test reactivated: 0.18% - non-reactivated: 0.14%; HOA dual-task reactivated: 0.15% - non-reactivated: 0.11%), and were excluded from the calculation of the median RT per block and per condition (reactivated vs. not-reactivated sequence). Accuracy of the key presses was used as secondary outcome, and it was calculated as the percentage of correct key presses per block and per condition, excluding the outliers identified in the RT data.

#### Analysis

We firstly investigated whether the two sequences showed a similar trend in performance improvement across initial acquisition. For this purpose, we used a Bayesian ANOVA including group (PD/HOA), condition (reactivated/non-reactivated) and block (10), and subject as random effect, and reported the BFs for each of these variables when compared to the null model see **Supplementary Material, Initial Encoding, Differences between finger tapping sequences pre-nap based on intervention**, for detailed results). Additionally, we tested for differences between the two sequences at initial encoding with a similar model, but this time using the within-factor sequence (A/B) rather than the factor condition (see **Supplementary Material, Initial Encoding, Differences between finger tapping sequences pre-nap based on sequence**). Given the weak to moderate evidence for no difference between sequences (RT: BF_10_ = 0.13; accuracy: BF_10_ = 0.76), their effect was not accounted for in the following analyses. Next, we analysed whether the practiced sequences during the sequential SRTT were learnt, as compared to the random version of the SRTT. For this purpose, the relative changes between the first 4 blocks of practice pre-nap (irrespective of the sequence) and last four blocks of test post-night were calculated. For the random SRTT, the relative change between the four blocks pre-nap and those post-night was calculated. These values were then compared with a Bayesian ANOVA including task (random SRTT/sequential SRTT), group, and subject as random effect (see **Supplementary Material, Motor Learning**, for detailed results).

The primary behavioural endpoint was the offline performance change, calculated for post-nap and post-night separately. This performance change was calculated as the difference of the first four blocks of sequential SRTT post-nap (for the post-nap offline changes) or post-night (post-night offline changes), relative to the last four blocks of test pre-nap. We verified that performance across the blocks of test pre-nap reached a plateau with no further within-session improvement, using a Bayesian ANOVA including group and block and their interaction as fixed effects, and subject as random effect. Here we found only weak evidence in favour of a difference across blocks (RT: BF_10_ = 2.27; accuracy: BF_10_ = 0.45), see **Supplementary Material, Performance Plateau**, for detailed results). Given the weak evidence, we opted to maintain all the blocks of test pre-nap for the calculation of the offline changes. The post-nap and post-night offline changes were then added to a Bayesian ANOVA, including group and condition (reactivated/non-reactivated), time (post-nap/post-night) and their interaction as fixed effects, and subject as random effect. The post-night offline changes, being relative to pre-nap performance, represent a sum of the changes related to the 2-hour nap with TMR, post-nap extended practice and overnight consolidation.

To evaluate whether a nap with TMR would have an effect on dual-tasking performance after the nap at 24-hour retention the dual-task cost was calculated as the relative difference of dual-task performance (two blocks) and single-task test performance, both at post-nap and post-night, respectively. These values were included in a Bayesian ANOVA including group, condition and session (post-nap/post-night) and their interactions as fixed effects, and subject as random effect. Details of the statistical analysis are reported in **Supplementary Material, Dual-task Results**.

We further performed explorations on the effects of TMR on extended practice, where we analysed the trend in performance across training post-nap and post-night, separately. For this purpose, two Bayesian ANOVA’s were used, with the same structure as that used for the analysis of initial encoding, using the data of the post-nap and post-night training blocks respectively (group x condition x block as fixed effects, subject as random effect). These analyses are reported in **Supplementary Material, Effects of Targeted Memory Reactivation on Extended Practice**.

### Electroencephalography

#### Offline sleep scoring

The 2-hour naps PSG recordings were post-hoc manually scored by a sleep researcher on 30-second epochs using the fMRI Artefact rejection and Sleep scoring Toolbox (FASST) (Leclercq et al., 2011), implemented in MATLAB, following the AASM guidelines (Berry et al., 2018). The researcher was not blinded for the group and the condition. Several metrics of sleep macro-architecture were extracted (i.e., total sleep time and time in bed, duration and percentage of each sleep stage, relative to total time in bed). Additionally, sleep efficiency was computed as the percentage of time spent in sleep (e.g., NREM1, NREM2, NREM3 and REM) relative to the total time in bed (for detailed descriptions of sleep macro-architecture, see Table 1.

#### Preprocessing

EEG data of NREM2 and NREM3 was selected for further preprocessing, performed using functions implemented in the FieldTrip Toolbox (Oostenveld et al., 2011) in MATLAB. First, EEG data were band-pass filtered (0.5-30Hz), using a low-order digital Butterworth filter. Next, data were manually cleaned from muscle activity and eye movement artifacts, using 30-s epoch screening. After artifact rejection, data were re-referenced to the average of A1-A2, and re-sampled to 500 Hz followed by Independent Component Analysis, to select and exclude components containing cardiac artifacts.

Micro-architecture metrics of sleep spindles (i.e., amplitude, frequency, absolute count), slow waves (i.e., amplitude, slope, absolute count) and their coupling were then extracted from the pre-processed data using YASA toolbox (Vallat & Walker, 2021) implemented in the Python environment. To extract spindles, we used the YASA spindle detection algorithm adapted from (Lacourse et al., 2019). Specifically, the relative power in the spindle frequency range (10–16Hz) was calculated as a ratio to the total power in the broad-band range (1–30Hz) using Short-Time Fourier Transforms with 2-second windows and a 200ms overlap. Next, the moving root mean squared (RMS) of the EEG data filtered in the sigma band was computed with a 300ms window with a 100ms step size, as well as a moving correlation between the broadband signal (1–30Hz) and the EEG signal filtered in the spindle frequency band. After these computation steps, spindle detection was bound to the simultaneous reaching of the following thresholds: (1) relative power in the sigma band (with respect to total power) above 0.2; (2) a moving RMS crossing the RMS mean + 1.5 RMS standard deviation threshold and (3) moving correlation above 0.65. We discarded any detected spindles with duration below 0.3s or above 3s, and we considered the same spindle and consequently merged events occurring on different channels within 500ms of each other.

The YASA slow wave detection algorithm was used to detect slow waves (Carrier et al., 2011; Massimini et al., 2004). Firstly, a bandpass filter between 0.3–1.5Hz using a Finite Impulse Response (FIR) filter was applied, and from the resulting signal the algorithm automatically detected any negative peak with an amplitude between –40 and –200μV and positive peaks with an amplitude comprised between 10–150μV. From these data, events constituted of a negative peak and the nearest following positive peak were defined as true slow waves if they fulfilled these criteria: (1) the negative deflection of duration 0.3–1.5 s, (2) positive deflection of duration 0.1–1.0s, (3) negative peak amplitude between 40–200 μV, (4) positive peak amplitude between 10–150μV and (5) peak-to-peak amplitude between 75– 350 μV. The coupling between the phase of the slow waves (0.3 – 1.5 Hz) and the amplitude of the sigma band of the spindles (10 – 16Hz) was computed as part of the slow wave detection algorithm. Specifically, the phase at sigma peak was extracted as the slow-wave phase at the point of maximal sigma amplitude within a 2-s epoch centred on the slow-wave negative peak, measured in radians.

Only the events identified as spindles and slow waves from channels C3 and Fz, respectively, were used for further statistical analysis. Of these events, spindle and slow wave amplitude, as well as spindle frequency were extracted, for the TMR and silent intervals separately (i.e. NREM 2–3 epochs without auditory stimulation). The phase at sigma peak was extracted from channel C3, where most spindles were detected. Density of spindles and slow waves was computed as the number of events detected in C3 and Fz, respectively, per percentage of cumulative NREM2-NREM3 sleep spent with TMR and with no-TMR, relative to the total time spent in NREM2 and NREM3 sleep. Values of detected spindles and slow-waves, relative to the cumulative time during the NREM2 and NREM3 sleep spent with TMR, are presented in Table 2.

**Table 2.** Description of the amount of NREM2-3 spent with auditory stimulation and in silence, and number of spindles and slow waves detected in these respective periods. Values are expressed as mean and range.

|  | PD | HOA |
| --- | --- | --- |
| NREM2-NREM3 with auditory stimulation (min) | 19.9<br>[1.6 – 45.6] | 17.1<br>[0.6 – 38.9] |
| NREM2-NREM3 in silence (min) | 36.9<br>[20.7 – 69.0] | 33.4<br>[1.4 – 61.6] |
| Spindles during auditory stimulation | 90.1<br>[2–248] | 83.4<br>[1–170] |
| Spindles during silence | 187<br>[9–388] | 185<br>[17–335] |
| Slow waves during auditory stimulation | 92<br>[1–520] | 59.7<br>[4–212] |
| Slow waves during silence | 152<br>[2–1119] | 89.7<br>[6–342] |

Electrophysiological data was extracted for 18 people with PD and 18 HOA. Spindles were detected for all these participants in both conditions. Notably, no slow waves were detected for one HOA and one person with PD, and one participant in the PD group did not present slow waves in the TMR period (stimulation: 17 people with PD and 17 HOA; silence: 16 people with PD and 17 HOA). For the phase at sigma peak computed at C3 values were obtained for 13 people with PD and 14 HOA in the TMR periods, and for 15 people with PD and 13 HOA in the silence periods, out of the 18 participants per group where these data were available. Missing values were excluded from the analyses.

Spindles and slow waves metrics were considered as non-physiological outliers if they fell above or below 3 standard deviations of each participant’s mean (spindle amplitude: PD – reactivated = 1.07%, non-reactivated = 1.07%; HOA – reactivated = 1.53 %, non-reactivated = 1.32 %; spindle frequency: PD – reactivated = 0.06%, non-reactivated = 0%; HOA reactivated = 0%, non-reactivated = 0%; slow wave amplitude: PD – reactivated = 2.05 %, non-reactivated = 2.41%; HOA – reactivated = 1.15%, non-reactivated = 1.88%) and were excluded from further analyses. Given the circular nature of the phase at sigma peak data, outliers were identified by firstly computing the circular mean and standard deviation with the “circular” package implemented in R (Agostinelli & Lund, 2023). Next, outliers were identified if lying 2 circular standard deviations from the circular mean, according to the von Mises distribution (Mardia & Jupp, 2000) (PD – reactivated = 1.40 %, non-reactivated = 0.94%; HOA – reactivated = 3.24%, non-reactivated = 2.70%).

#### Analysis

The extracted sleep macro- and micro-architecture metrics (spindle density, amplitude, frequency, slow wave density, amplitude) were compared using Bayesian ANOVAs, including group and condition (periods of TMR/periods of no-TMR) as fixed effects, and subject as random effect. Findings on spindle frequency are reported in **Supplementary Material, The Modulatory effect of TMR on Spindle Frequency**.

To study the phase at sigma peak during TMR and during silence, the angular mean of the phase at sigma peak during the two conditions was calculated. If data followed the Von Mises distribution, tested with the Rayleigh test for non-uniformity (p > 0.05), the Watson–Williams multi-sample test for equal means was used, including the mean phase at sigma peak of each subject and the group (PD/HOA), for TMR and silence periods separately, using the ‘CircStats’ package in R (Lund Ulric & Agostinelli Claudio, 2025). Conversely, if the assumption was not met, the Watson’s U^2^ test for homogeneity of two samples of circular data was implemented from the “circular” package, with alpha = 0.05. Additionally, the Watson’s U^2^ test was used to compare within-group differences between TMR periods and silence periods. Results of the Watson’s U^2^ test are reported as test statistic, critical value for the defined alpha and p-value range.

We also explored the relationship between sleep metrics and behavioural performance, specifically offline changes post-intervention, accounting for the group, and the condition (here intended as the periods with or without TMR during the 2-hour nap opportunity) using Bayesian linear mixed models. For this specific analysis, we modelled the sleep micro-architecture data from the TMR intervals, and we used it, together with the group factor and the interaction between group and sleep metric, as a predictor of behavioural performance of the reactivated and non-reactivated sequence separately. In case of at least moderate evidence for an effect, we performed a post-hoc analysis to study the contrasts and compare the slopes using the R package “emmeans” package (Russell V. Lenth, 2023).

The “circlin.cor” function from the Directional package (Mardia & Jupp, 2000; Tsagris Michail et al., 2025) was implemented, computing the squared correlation between the phase at sigma peak and the post-nap offline changes. Results of this analysis were reported as R^2^ and p-values, and FDR-correction for multiple comparisons was used.

## Data Availability

All data produced in the present study are available upon reasonable request to the authors

## Acknowledgements

This project was funded by the Funds Malou Malou & Perano, managed by the King Baudouin Foundation (2019-J4121350-212854) and Internal Funds KU Leuven (STG/21/035). Moran Gilat was supported by a Marie-Skłodowska-Curie Action Postdoctoral Fellowship (TARGET-SLEEP, 838576). Letizia Micca is supported by a FWO PhD Fellowship (SLEEP ON IT, 11PQB24N). Judith Nicolas was funded by The Fondation Pour la Recherche Médicale (ARF202309017485). The funders played no role in study design, data collection, analysis and interpretation of data, or the writing of this manuscript.

The authors would like to thank Dr Menno Veldman for his assistance with the protocol development.

## Data availability

The datasets generated and/or analysed during the current study are available upon reasonable request and provision of ethical approval for re-use of the data and data sharing agreement.

## Code availability

The underlying code for this study is available upon request.

