## Supplementary material for "Targeted memory reactivation during sleep modulates spindle and slow wave density, but not motor memory consolidation, in Parkinson’s disease"

### Psychomotor Vigilance Test

Table 1. Data of the Psychomotor Vigilance Test (PVT) across sessions, presented as mean and 95% CI. Note that for the post-night session data of one person with PD is missing, and data of one HOA is missing for all the sessions, thus data of these participants was excluded from the calculations. Lapses are defined as values above 500ms.
Parkinson’s disease – PD, and healthy older adults – HOA

|  |  | Pre-nap | Post-nap | Post-night |
| --- | --- | --- | --- | --- |
| PD = 19 | RT | 0.36 [0.33 – 0.39] | 0.34 [0.32 – 0.37] | 0.37 [0.33 – 0.40] |
|  | Lapses | 12.7 [6.1 – 19.4] | 9 [3.5 – 14.5] | 13.8 [4.7 – 22.9] |
|  | Fastest 10% | 0.26 [0.25 – 0.27] | 0.25 [0.23 – 0.27] | 0.27 [0.25 – 0.28] |
|  | Slowest 10% | 0.77 [0.59 – 0.95] | 0.59 [0.49 – 0.69] | 0.64 [0.51 – 0.76] |
| HOA = 19 | RT | 0.33 [0.32 – 0.35] | 0.33 [0.31 – 0.34] | 0.33 [0.31 – 0.34] |
|  | Lapses | 5.7 [3.5 – 7.9] | 5 [2.9 – 7.1] | 3.5 [1.3– 5.6] |
|  | Fastest 10% | 0.27 [0.25 – 0.28] | 0.26 [0.25 – 0.27] | 0.27 [0.26 – 0.28] |
|  | Slowest 10% | 0.54 [0.48 – 0.59] | 0.53 [0.49 – 0.57] | 0.49 [0.46 – 0.51] |

Reaction time values of the Psychomotor Vigilance Test (PVT) were firstly screened for outliers using the Tukey method. Namely, values were screened for each subject and session separately, and they were excluded if they lied below or above 1.5 interquartile range. The median of the values for each subject and session was then calculated and used for further analyses (see Table 1 for detailed descriptives).

Analysis of the PVT was performed on the median reaction time of the key presses with a Bayesian ANOVA including group (PD, HOA), time (pre-nap, post-nap and post-night) and their interaction as fixed effects and subject as random effect. Note that for one HOA group data of the task was missing due to a technical issue during the experimental procedures, and one person with PD did not perform the PVT at post-night for technical issues. Data of these were excluded from the analysis. Findings revealed weak evidence for all the main effects and interactions (group: BF_10_ = 2.11, F_(1,36)_ = 3.48, p = 0.07, ges = 0.08; intervention: BF_10_ = 1.45, F_(2,72)_ = 4.23, p = 0.02, p(GG) = 0.03, ges = 0.01; group by time: BF_10_ = 2.11, F_(2,72)_ = 4.13, p = 0.02, p(GG) = 0.03, ges = 0.01).

### Motor Learning

Reaction time. To assess whether motor performance of the trained sequences, measured with reaction time (RT), benefitted from training compared to a random sequence, we calculated the overall change of performance at post-night, relative to pre-test, of the trained sequences and the random sequences. Subsequently, we performed a Bayesian ANOVA including group, task (trained serial reaction time task – SRTT, random sequence) as fixed effects, and subject as random effect. The random sequence relative change between pre-nap and post-night differed from that of the sequences trained, as confirmed by the decisive level of evidence of the Bayesian ANOVA (BF_10_ > 100, F_(1,38)_ = 63.77, p < 0.01, ges = 0.35). Weak level of evidence was found for no main effect of group (BF_10_ = 0.55, F_(1,38)_ = 1.89, p = 0.18, ges = 0.03) and for no group by task interaction (BF_10_ = 0.38, F_(1,38)_ = 0.69, p = 0.41, ges < 0.01).

A similar model was implemented to study whether participants presented differences in the performance of the sequential SRTT and the random SRTT between pre and post-nap. We compared performance of the first 4 blocks of sequential SRTT with the random SRTT pre-nap, finding moderate evidence for a difference between tasks (BF_10_ = 6.73, F_(1,38)_ = 8.41, p < 0.01, ges < 0.01) but weak evidence for a difference between groups or an interaction effect (group: BF_10_ = 0.57, F_(1,38)_ = 1.48, p = 0.30, ges = 0.04; group by task: BF_10_ = 0.47, F_(1,38)_ = 1.33, p = 0.26, ges < 0.01).

Sequential SRTT performance of retest and random SRTT post-nap were also compared, finding decisive evidence for a task difference (BF_10_ > 100, F_(1,38)_ = 43.36, p < 0.01, ges = 0.05), weak evidence for a group difference (BF_10_ = 1.10, F_(1,38)_ = 2.16, p = 0.15, ges = 0.05), and moderate evidence for no interaction effect (BF_10_ = 0.31, F_(1,38)_ = 0.25, p = 0.62, ges < 0.01).

Finally, performance of the sequential SRTT at retest and random SRTT post-night were compared, showing decisive evidence for a task effect (BF_10_ > 100, F_(1,38)_ = 94.16, p < 0.01, ges = 0.13), weak evidence for a difference between groups (BF_10_ = 1.04, F_(1,38)_ = 2.72, p = 0.11, ges = 0.06) and no interaction effect (BF_10_ > 100, F_(1,38)_ = 94.16, p < 0.01, ges = 0.13).

Accuracy. We found strong evidence for an effect of task (BF_10_ = 18.38, F_(1,38)_ = 12.42, p < 0.01, ges = 0.13), and moderate evidence for no effect of group (BF_10_ = 0.24, F_(1,38)_ = 0.15, p = 0.70, ges < 0.01) or group by task interaction (BF_10_ = 0.27, F_(1,38)_ = 0.001, p = 0.97, ges < 0.01).

Comparisons between average initial performance of the sequential SRTT and random SRTT pre-nap did not show any differences between groups and tasks, nor an interaction effect (group: BF_10_ = 0.47, F_(1,38)_ = 0.009, p = 0.92, ges < 0.01; task: BF_10_ = 0.23, F_(1,38)_ = 0.03, p = 0.87, ges < 0.01; group by task: BF_10_ = 0.32, F_(1,38)_ = 0.11, p = 0.74, ges < 0.01).

Task differences were highlighted post-nap and post-night (post-nap: BF_10_ = 5.88, F_(1,38)_ = 7.93, p < 0.01, ges = 0.03; post-night: BF_10_ > 100, F_(1,38)_ = 18.97, p < 0.01, ges = 0.07), but group differences or interactions were not evidenced in neither of these sessions (post-nap – group: BF_10_ = 0.80, F_(1,38)_ = 1.68, p = 0.20, ges = 0.04; group by task: BF_10_ = 0.30 F_(1,38)_ = 0.03, p = 0.91, ges < 0.01; post-night – group: BF_10_ = 0.38, F_(1,38)_ = 0.02, p = 0.88, ges < 0.01; group by task: BF_10_ = 0.31, F_(1,38)_ = 0.13, p = 0.72, ges < 0.01).

The findings suggest a difference in reaction time and accuracy change between trained and random tasks, which follows a similar trend both in people with PD and HOA and suggest that improvement in performance was not due to mere motor execution.

### Initial Encoding

#### Differences between finger tapping sequences pre-nap based on condition

Reaction time. Analysis of performance at initial training was conducted with a Bayesian ANOVA including group, condition (reactivated, non-reactivated), and block (10). Results showed strong evidence for a group by block interaction (BF_10_ = 17.65, F_(9,342)_ = 2.97, p < 0.01, p(GG) = 0.03 ges < 0.01) and decisive evidence for an effect of block (BF_10_ > 100, F_(9,342)_ = 8.22, p < 0.01, p(GG) < 0.01, ges = 0.01). Weak to moderate evidence was found for no effect of group and condition (group: BF_10_ = 0.76, F_(1,38)_ = 2.32, p = 0.14, ges = 0.05; condition: BF_10_ = 0.28, F_(1,38)_ = 1.73, p = 0.19, ges < 0.01), and the other interactions showed moderate to strong evidence for no effect (group by condition: BF_10_ = 0.20, F_(1,38)_ = 0.88, p = 0.35, ges < 0.01; condition by block: BF_10_ = 0.007, F_(9,342)_ = 1.24, p = 0.27, p(GG) = 0.29, ges < 0.01; group by condition by block: BF_10_ = 0.02, F_(9,342)_ = 0.95, p = 0.48, p(GG) = 0.45, ges < 0.01).

Accuracy. Results showed overall moderate to strong evidence for no main effect or interaction (group: BF_10_ = 0.30, F_(1,38)_ = 0.14, p = 0.71, ges < 0.01; condition: BF_10_ = 0.13, F_(1,38)_ = 0.40, p = 0.53, ges < 0.01; block: BF_10_ = 0.01, F_(9,342)_ = 1.48, p = 0.15, p(GG) = 0.18, ges < 0.01; group by condition interaction: BF_10_ = 0.15, F_(1,38)_ = 0.25, p = 0.62, ges < 0.01; group by block interaction: BF_10_ = 0.003, F_(9,342)_ = 0.48, p = 0.89, p(GG) = 0.83, ges < 0.01 condition by block interaction: BF_10_ = 0.02, F_(9,342)_ = 1.21, p = 0.29, p(GG) = 0.30, ges < 0.01; group by condition by block interaction: BF_10_ = 0.02, F_(9,342)_ = 0.65, p = 0.76, p(GG) = 0.71, ges < 0.01).

While the findings on reaction time indicate the existence of a baseline difference between people with PD and HOA in the performance improvement during initial training, performance accuracy in both groups did not vary across initial training.

#### Differences between finger tapping sequences pre-nap based on sequence

Reaction time. Performance of the two sequences during initial learning pre-nap was compared using a Bayesian ANOVA, including group (PD, HOA), sequence (A/B) and practice block (10) as fixed effects, and subject as random effect. We found strong evidence for a group by block interaction (BF_10_ = 17.67, F_(9,342)_ = 2.97, p < 0.01, p(GG) = 0.03, ges < 0.01) and decisive evidence for an effect of block (BF_10_ > 100, F_(9,342)_ = 8.22, p < 0.01, p(GG) < 0.01, ges = 0.01). Conversely, there was weak to moderate evidence for no main effect of group (BF_10_ = 0.98, F_(1,38)_ = 2.32, p = 0.14, ges = 0.05), sequence (BF_10_ = 0.13, F_(1,38)_ = 0.60, p = 0.44, ges < 0.01), or for the group by sequence interaction (BF_10_ = 0.26, F_(1,38)_ = 1.16, p = 0.29, ges < 0.01), and strong evidence for no effect of the sequence by block interaction (BF_10_ = 0.003, F_(9,342)_ = 0.83, p = 0.59, p(GG) = 0.54, ges < 0.01). The three-way interaction of group by sequence by block also showed moderate evidence for no effect (BF_10_ = 0.02, F_(9,342)_ = 0.94, p = 0.49, p(GG) = 0.46, ges < 0.01).

Accuracy. The findings of this analysis highlighted no effect of any of the main factors and interactions, with a level of evidence varying from weak to strong (group: BF_10_ = 0.29, F_(1,38)_ = 0.14, p = 0.71, ges < 0.01; sequence: BF_10_ = 0.76, F_(1,38)_ = 1.97, p = 0.17, ges < 0.01; block: BF_10_ = 0.01, F_(9,342)_ = 1.48, p = 0.15, p(GG) = 0.18, ges < 0.01; group by sequence interaction: BF_10_ = 0.22, F_(1,38)_ = 0.60, p = 0.44, ges < 0.01; group by block interaction: BF_10_ = 0.003, F_(9,342)_ = 0.48, p = 0.89, p(GG) = 0.83, ges < 0.01 sequence by block interaction: BF_10_ = 0.006, F_(9,342)_ = 0.79, p = 0.63, p(GG) = 0.60, ges < 0.01; group by sequence by block interaction: BF_10_ = 0.06, F_(9,342)_ = 1.21, p = 0.29, p(GG) = 0.30, ges < 0.01).

Overall, the findings on reaction time are indicative of a performance change across initial training, which is similar between the sequences. However, the group by block interaction hints to a difference in improvement trend between PD and HOA. Conversely, the findings on accuracy did not highlight differences between the sequences trained before the nap, nor a difference between groups. Because of the lack of difference between sequences, the factor sequence was not further included in any model.

### Performance Plateau

Reaction time. To test whether performance plateau was reached at test pre-nap, a Bayesian ANOVA group and block (4, irrespective of the sequence practiced) and their interaction as fixed effects, and subject as random effect. The results showed weak evidence in favour of an effect of block (BF_10_ = 2.27, F_(3,114)_ = 3.58, p < 0.01, p(GG) = 0.02, ges < 0.01). and for an effect of group (BF_10_ = 1.10, F_(1,38)_ = 2.40, p = 0.13, ges = 0.06). Moderate evidence for no group by block interaction was found (BF_10_ = 0.11, F_(3,114)_ = 0.44, p = 0.72, p(GG) = 0.69, ges < 0.01).

Accuracy. Results on pre-nap test performance showed weak to moderate evidence for no effect of group (BF_10_ = 0.30, F_(1,38)_ = 0.16, p = 0.69, ges < 0.01), block (BF_10_ = 0.45, F_(3,114)_ = 2.17, p = 0.10, p(GG) = 0.11, ges = 0.02) nor their interaction (BF_10_ = 0.13, F_(3,114)_ = 0.66, p = 0.58, p(GG) = 0.56, ges < 0.01).

These findings suggest that performance, measured both with RT and with accuracy, was stable across the blocks of test pre-nap, irrespective of the group or the condition.

### Dual-task Results

Reaction time. Weak to moderate evidence for no main effects or interactions was found for RT (group: BF_10_ = 0.28, F_(1,38)_ = 0.10, p = 0.75, ges < 0.01; condition: BF_10_ = 0.17, F_(1,38)_ = 0.007, p = 0.94, ges < 0.01; time: BF_10_ = 0.17, F_(1,38)_ = 0.08, p = 0.78, ges < 0.01; group by condition: BF_10_ = 0.23, F_(1,38)_ = 0.003, p = 0.95, ges < 0.01; group by time: BF_10_ = 0.39, F_(1,38)_ = 1.08, p = 0.31, ges < 0.01; condition by time: BF_10_ = 0.33, F_(1,38)_ = 0.92, p = 0.34, ges < 0.01; group by condition by time: BF_10_ = 0.34, F_(1,38)_ = 0.41, p = 0.53, ges < 0.01, see Figure 1).

Accuracy. The results showed overall weak to moderate evidence for no effect of any of the main factors and interactions (group: BF_10_ = 0.27, F_(1,38)_ = 0.96, p = 0.33, ges < 0.01; condition: BF_10_ = 0.33, F_(1,38)_ = 1.24, p = 0.27, ges < 0.01; time: BF_10_ = 0.18, F_(1,38)_ = 0.11, p = 0.74, ges < 0.01; group by condition interaction: BF_10_ = 0.82, F_(1,38)_ = 2.39, p = 0.13, ges = 0.02; group by time interaction: BF_10_ = 0.45, F_(1,38)_ = 1.89, p = 0.18, ges < 0.01; condition by time interaction: BF_10_ = 0.38, F_(1,38)_ = 0.78, p = 0.38, ges < 0.01; group by condition by time interaction: BF_10_ = 0.34, F_(1,38)_ = 0.16, p = 0.69, ges < 0.01; see Figure 2).

Figure 1. Dual-task costs, measured with reaction time, whereby lower values indicate better performance on the dual-task. Violin plot: mean (diamond), median (central horizontal bar), and 25^th^ (lower bar) and 75^th^ (higher bar) percentiles. DT: Dual-task

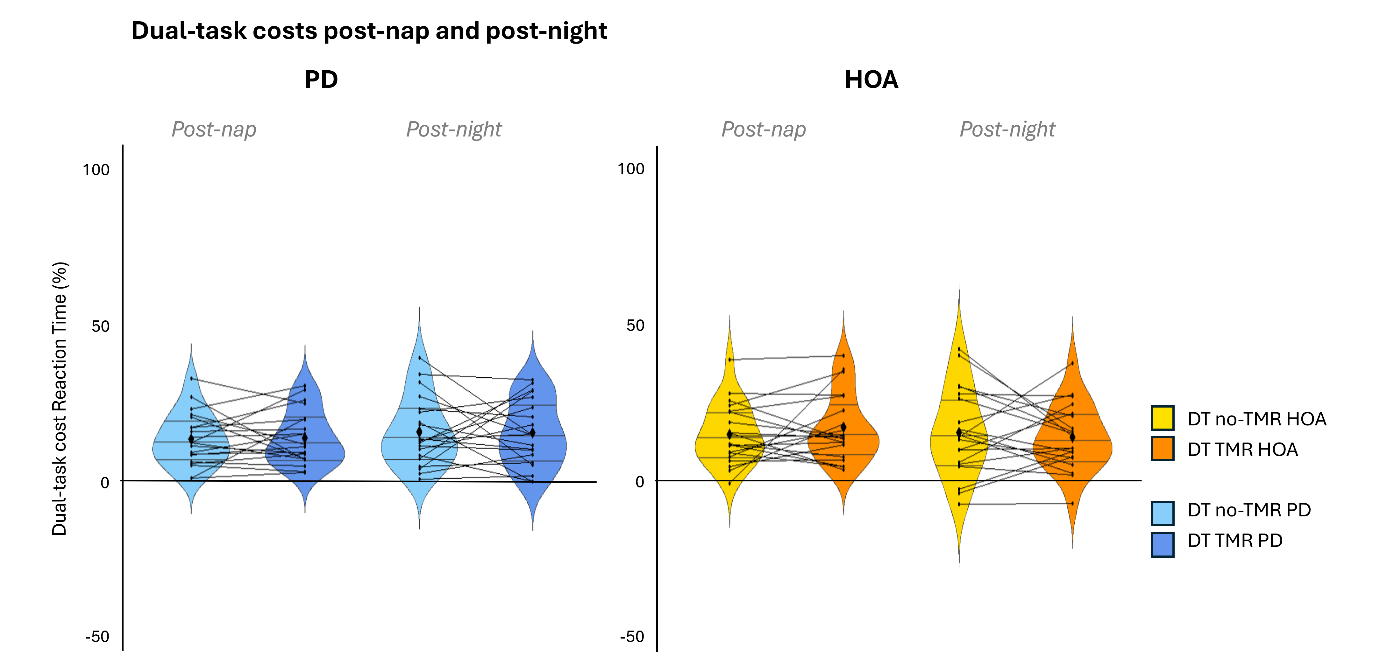

Figure 2. Dual-task costs, measured with accuracy, whereby lower values indicate better performance on the dual-task. Violin plot: mean (diamond), median (central horizontal bar), and 25^th^ (lower bar) and 75^th^ (higher bar) percentiles. DT: Dual-task

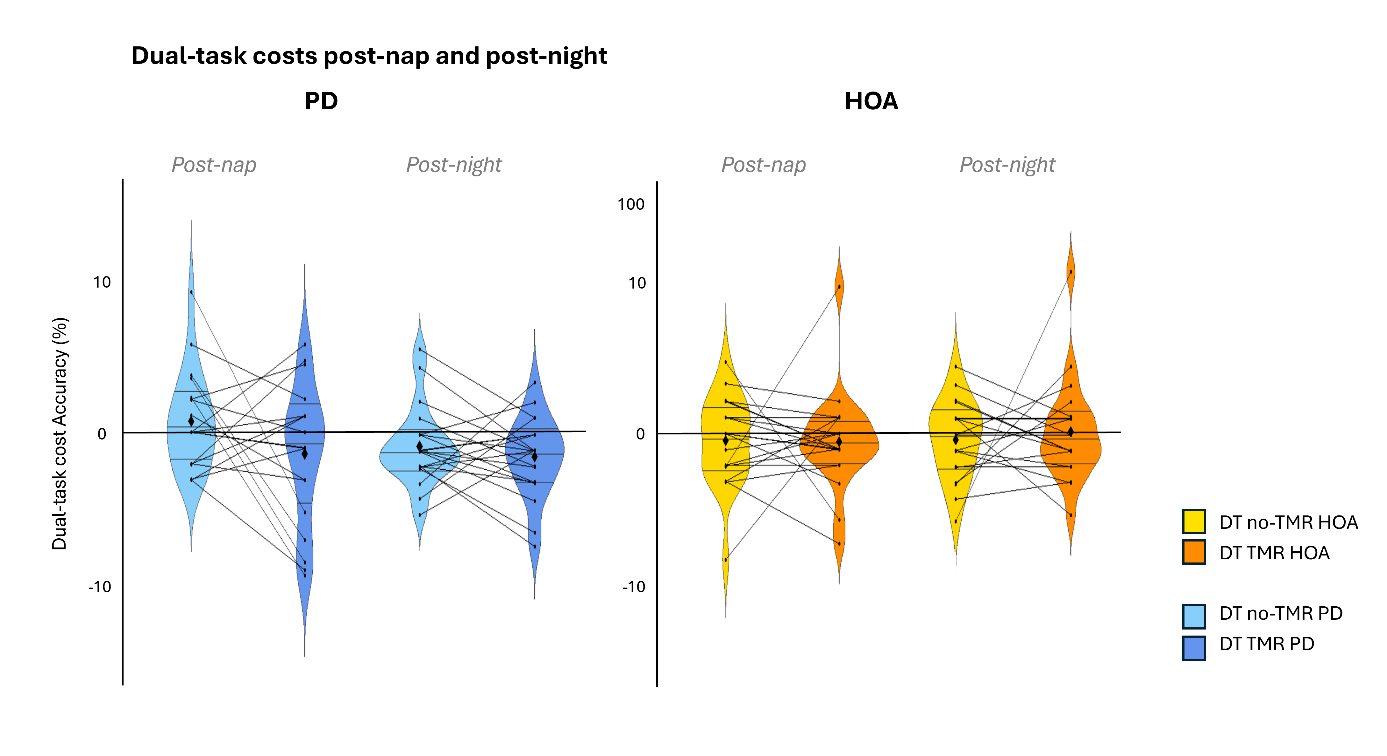

### Effects of Targeted Memory Reactivation on Extended Practice

Reaction time. The performance trend post-nap and post-night was modelled with two separate Bayesian ANOVAs, including group, condition and block as fixed effects, and subject as random effect. At post-nap, we found decisive evidence for an effect of block (BF_10_ > 100, F_(9,342)_ = 11.64, p < 0.01, p(GG) < 0.01, ges < 0.01). The evidence for no effect of group was weak (BF_10_ = 0.73, F_(1,38)_ = 2.04, p = 0.16, ges = 0.05), and it was moderate to strong in favour of no effect of the condition and of the interactions (condition: BF_10_ = 0.09, F_(1,38)_ = 0.07, p = 0.79, ges < 0.01; group by condition: BF_10_ = 0.14, F_(1,38)_ = 0.14, p = 0.71, ges < 0.01; group by block: BF_10_ = 0.008, F_(9,342)_ = 1.02, p = 0.42, p(GG) = 0.41, ges < 0.01; condition by block: BF_10_ = 0.006, F_(9,342)_ = 1.03, p = 0.41, p(GG) = 0.40, ges < 0.01; group by condition by block: BF_10_ = 0.01, F_(9,342)_= 0.64, p = 0.76, p(GG) = 0.69, ges < 0.01).

Similarly, at post-night there was decisive evidence for an effect of block (BF_10_ > 100, F_(9,342)_ = 9.27, p < 0.01, p(GG) < 0.01, ges < 0.01), but here it was weakly favouring no effect of group (BF_10_ = 0.87, F_(1,38)_ = 2.80, p = 0.10, ges = 0.06) and condition (BF_10_ = 0.42, F_(1,38)_ = 0.51, p = 0.48, ges < 0.01), and it was moderately to strongly in favour of no interaction effect (group by condition: BF_10_ = 0.12, F_(1,38)_ = 0.03, p = 0.86, ges < 0.01; group by block: BF_10_ = 0.003, F_(9,342)_ = 0.58, p = 0.82, p(GG) = 0.68, ges < 0.01; condition by block: BF_10_ = 0.002, F_(9,342)_ = 0.82, p = 0.60, p(GG) = 0.56, ges < 0.01; group by condition by block: BF_10_ = 0.01, F_(9,342)_= 0.84, p = 0.58, p(GG) = 0.55, ges < 0.01).

Accuracy. At post-nap, we found strong evidence for an effect of block (BF_10_ = 14.93, F_(9,342)_ = 3.37, p < 0.01, p(GG) < 0.01, ges = 0.02). There was weak evidence for no effect of group (BF_10_ = 0.42, F_(1,38)_ = 1.06, p = 0.31, ges = 0.01), while this was moderately to strongly in favour of no effect of condition (BF_10_ = 0.08, F_(1,38)_ = 0.0003, p = 0.99, ges < 0.01), group by condition interaction (BF_10_ = 0.12, F_(1,38)_ = 0.05, p = 0.83, ges < 0.01), group by block interaction (BF_10_ = 0.03, F_(9,342)_ = 1.27, p = 0.25, ges < 0.01) or group by condition by block interaction (BF_10_ = 0.24, F_(9,342)_ = 1.89, p = 0.05, ges = 0.01), and it was strongly in favour of no group by condition interaction (BF_10_ = 0.003, F_(9,342)_ = 0.54, p = 0.84, ges < 0.01).

At post-night, we found weak evidence for no effect of group (BF_10_ = 0.33, F_(1,38)_ = 0.90, p = 0.35, ges < 0.01), and moderate to strong evidence for no other main effect nor interaction (condition: BF_10_ = 0.32, F_(1,38)_ = 1.77, p = 0.19, ges < 0.01; block: BF_10_ = 0.001, F_(9,342)_ = 0.74, p = 0.67, p(GG) = 0.62, ges < 0.01; group by condition interaction: BF_10_ = 0.15, F_(1,38)_ = 0.37, p = 0.54, ges < 0.01; group by block interaction: BF_10_ = 0.04, F_(9,342)_ = 1.33, p = 0.22, p(GG) = 0.25, ges = 0.01; condition by block interaction: BF_10_ = 0.09, F_(9,342)_ = 1.75, p = 0.08, p(GG) = 0.11, ges = 0.01; group by condition by block interaction: BF_10_ = 0.02, F_(9,342)_ = 0.68, p = 0.73, ges < 0.01).

These findings suggest no specific effect of TMR on reaction time and accuracy on the practice during post-nap and post-night sessions. However, reaction time and accuracy improvements could still be observed during online practice post-nap, while at post-night these were only evident when measured with reaction time.

### Stimulations per Sleep Stage

Table 2. Amount of stimulations (complete 8-tone sequences) replayed during each sleep stage. Note that of the HOA, 3 presented 0 stimulations in wake, 5 had 0 stimulations in NREM3, 12 had 0 stimulations in REM. Of the people with PD, 2 presented 0 stimulations during wake, 3 showed 0 stimulations in NREM3 and 17 had 0 stimulations in REM.
Variables in absolute number and percentage are presented as the mean (±95% confidence interval), unless otherwise specified. Statistics were performed on the absolute number, the percentages are reported for completeness.
* significant p values according to frequentist statistics

|  | PD = 19 | HOA = 18 | Test result |
| --- | --- | --- | --- |
| Total | 210.5 [167.9 – 253.0] | 209.1 [161.2 – 256.9] | BF_10_ = 0.32,  t_(34)_ = 0.04, p = 0.96 |
|  | 100% | 100% |  |
| Wake | 7.7 [3.9 – 11.5] | 6.9 [2.4 – 11.5] | BF_10_ = 0.33,  W = 193, p = 0.51 |
|  | 4.54%  [1.17 – 7.90] | 4.46%  [1.32 – 7.60] |  |
| NREM1 | 14.8 [6.2 – 23.4] | 22.1 [14.6 – 29.5] | BF_10_ = 0.58,  W = 102.5, p = 0.04* |
|  | 7.58%  [3.46 – 11.68] | 16.35%  [8.18 – 24.53] |  |
| NREM2 | 109.2 [77.3 – 141.2] | 109.1 [83.2 – 135.0] | BF_10_ = 0.32,  W = 154, p = 0.61 |
|  | 55.21%  [43.12 – 67.29] | 52.42%  [44.07 – 60.76] |  |
| NREM3 | 75.9 [35.1 – 116.7] | 66.3 [32.4 – 100.2] | BF_10_ = 0.33,  W = 178.5, p = 0.83 |
|  | 30.99%  [17.86 – 44.12] | 24.75%  [13.93 – 35.57] |  |
| REM | 0.84 [0 – 2.1] | 2.5 [0 – 6.3] | BF_10_ = 0.41,  W = 136, p = 0.15 |
|  | 0.36%  [0.00 – 0.86] | 0.81%  [0.00 – 1.87] |  |

### The Modulatory Effects of TMR on Spindle Frequency

When comparing spindle frequency between periods with TMR cues and silent periods within the 2-hour nap, we found weak to moderate evidence for no effect of group (BF_10_ = 0.67, F_(1,34)_ = 0.90, p = 0.35, ges = 0.02), condition (BF_10_ = 0.88, F_(1,34)_ = 3.06, p = 0.09, ges < 0.01) and their interaction (BF_10_ = 0.32, F_(1,34)_ = 0.01, p = 0.93, ges < 0.01) (see Figure 1.A for details).

Figure 1. Spindle frequency during the auditory stimulation and silent periods.
Violin plot: mean (diamond), median (central horizontal bar), and 25^th^ (lower bar) and 75^th^ (higher bar) percentiles.

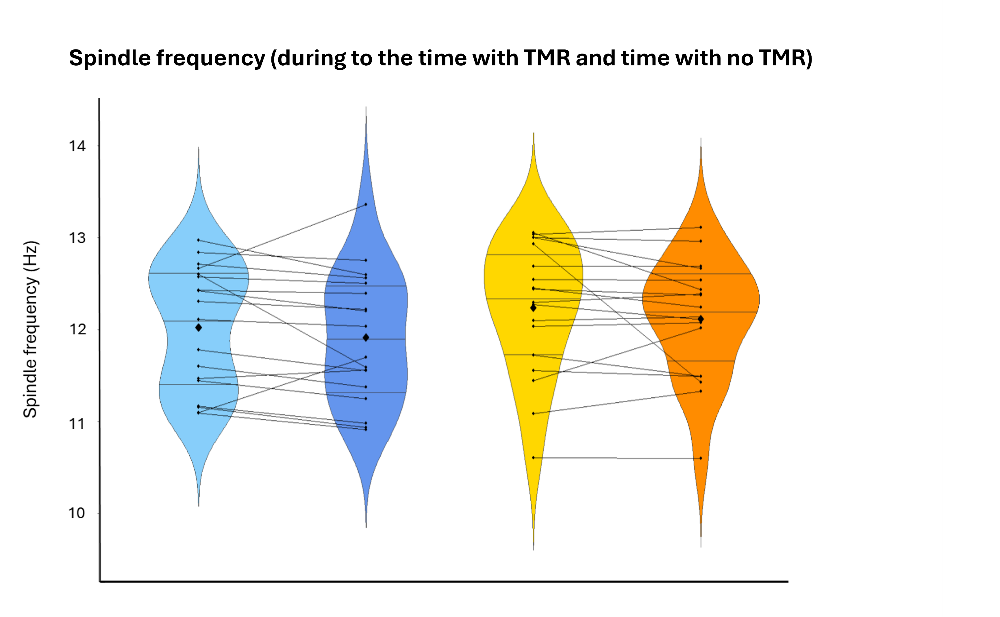

### Association between Spindle and Slow Wave Characteristics and Behaviour

#### Spindle density on non-reactivated sequence

Reaction time. The linear model used for this analysis highlighted weak evidence for no effect of group (BF_10_ = 0.33, F_(1)_ = 0.66, p = 0.42, ges = 0.02), spindle density during TMR periods (BF_10_ = 0.34, F_(1)_ = 0.95, p = 0.34, ges = 0.03) or their interaction (BF_10_ = 0.47, F_(1)_ = 0.98, p = 0.33, ges = 0.03) on the offline changes of the non-reactivated sequence measured with reaction time.

Accuracy. Similarly, weak to moderate evidence was found for no effect of group (BF_10_ = 0.49, F_(1)_ = 0.13, p = 0.72, ges < 0.01), spindle density during TMR periods (BF_10_ = 0.32, F_(1)_ = 0.11, p = 0.92, ges < 0.01) or their interaction (BF_10_ = 0.32, F_(1)_ = 0.11, p = 0.92, ges < 0.01) on the offline changes of the non-reactivated sequence measured with accuracy.

#### Slow wave density on non-reactivated sequence

Reaction time. The analysis of the offline changes of the non-reactivated sequence measured with reaction time showed weak evidence for no effect of group (BF_10_ = 0.34, F_(1)_ = 1.11, p = 0.30, ges = 0.03), slow wave density during TMR periods (BF_10_ = 0.35, F_(1)_ = 2.81, p = 0.10, ges = 0.07) or their interaction (BF_10_ = 0.61, F_(1)_ = 2.86, p = 0.10, ges = 0.07).

Accuracy. Similarly, weak evidence was found for no effect of group (BF_10_ = 0.40, F_(1)_ = 0.42, p = 0.53, ges = 0.01), slow wave density during TMR periods (BF_10_ = 0.34, F_(1)_ = 0.004, p = 0.95, ges < 0.01) or their interaction (BF_10_ = 0.35, F_(1)_ = 0.05, p = 0.83, ges < 0.01) on the offline changes of the non-reactivated sequence measured with accuracy.

#### Spindle amplitude on non-reactivated sequence

Reaction time. We found weak evidence for no effect of group (BF_10_ = 0.33, F_(1)_ = 0.79, p = 0.38, ges = 0.02), spindle amplitude during TMR periods (BF_10_ = 0.56, F_(1)_ = 2.10, p = 0.16, ges = 0.05) or their interaction (BF_10_ = 0.47, F_(1)_ = 0.75, p = 0.39, ges = 0.02) on the offline changes of the non-reactivated sequence measured with reaction time.

Accuracy. Weak evidence was found for no effect of group (BF_10_ = 0.49, F_(1)_ = 1.04, p = 0.32, ges = 0.03), spindle amplitude during TMR periods (BF_10_ = 0.35, F_(1)_ = 0.74, p = 0.40, ges = 0.02) or their interaction (BF_10_ = 0.42, F_(1)_ = 0.63, p = 0.43, ges = 0.02) on the offline changes of the non-reactivated sequence measured with accuracy.

#### Slow wave amplitude on non-reactivated sequence

Reaction time. The analysis showed weak evidence for no effect of group (BF_10_ = 0.34, F_(1)_ = 1.57, p = 0.22, ges = 0.05), slow wave amplitude during TMR periods (BF_10_ = 0.34, F_(1)_ = 0.78, p = 0.39, ges = 0.02) or their interaction (BF_10_ = 0.66 F_(1)_ = 1.67, p = 0.21, ges = 0.05) on the offline changes of the non-reactivated sequence measured with reaction time.

Accuracy. Weak evidence was also found for no effect of group (BF_10_ = 0.40, F_(1)_ = 0.08, p = 0.78, ges < 0.01), spindle amplitude during TMR periods (BF_10_ = 0.41, F_(1)_ = 0.50, p = 0.49, ges = 0.02) or their interaction (BF_10_ = 0.34, F_(1)_ = 0.14, p = 0.71, ges < 0.01) on the offline changes of the non-reactivated sequence measured with accuracy.

#### Spindle frequency

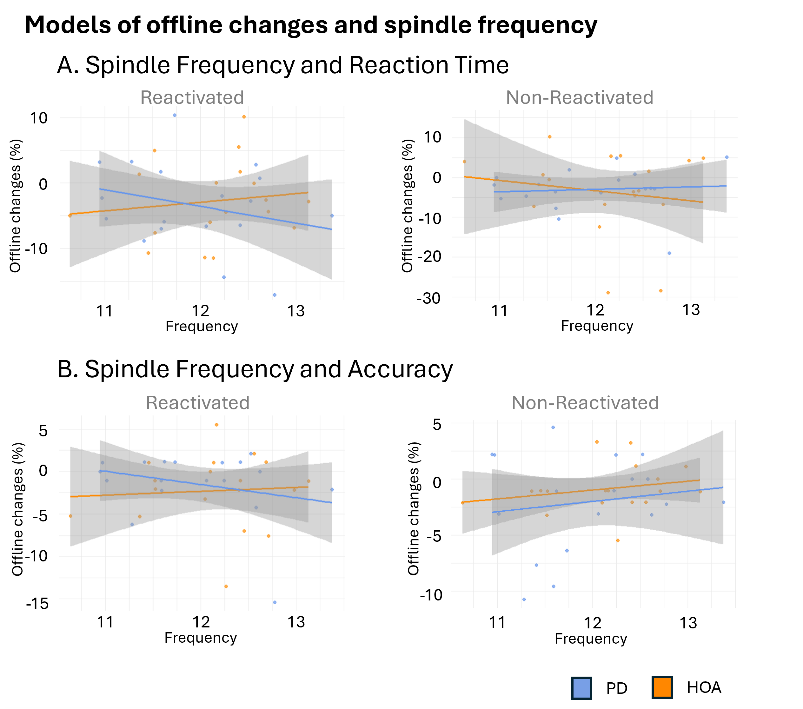
Reaction time. The linear model used for this analysis highlighted weak evidence for no effect of group (BF_10_ = 0.33, F_(1)_ = 1.34, p = 0.26, ges = 0.04), spindle frequency during TMR periods (BF_10_ = 0.35, F_(1)_ = 0.29, p = 0.59, ges < 0.01) or their interaction (BF_10_ = 0.59, F_(1)_ = 1.39, p = 0.25, ges = 0.04) on the offline changes of the reactivated sequence measured with reaction time. Analysis of the non-reactivated sequence showed weak to moderate evidence for no main effect or interaction (group: BF_10_ = 0.33, F_(1)_ = 0.48, p = 0.49, ges = 0.01; spindle frequency during TMR periods: BF_10_ = 0.34, F_(1)_ = 0.59, p = 0.45, ges = 0.02; group by spindle frequency: BF_10_ = 0.39, F_(1)_ = 0.50, p = 0.49, ges = 0.01) (see Figure 2.A for details).

Figure 2. Linear mixed models for the prediction of offline relative changes post-intervention with sleep micro-architecture metrics in PD (blue) and HOA (orange). (A) Spindle frequency and offline changes measured with reaction time, (B) and accuracy.

Dots represent individual values, with linear regression lines for PD and HOA. The grey areas indicate the standard error.

Accuracy. Weak evidence was found for no effect of group (BF_10_ = 0.37, F_(1)_ = 0.89, p = 0.35, ges = 0.03), spindle frequency during TMR periods (BF_10_ = 0.39, F_(1)_ = 0.08, p = 0.78, ges < 0.01) or their interaction (BF_10_ = 0.47, F_(1)_ = 0.84, p = 0.37, ges = 0.02) on the offline changes of the reactivated sequence measured with accuracy. Similar findings were highlighted by the analysis of the non-reactivated sequence (group: BF_10_ = 0.49, F_(1)_ = 0.01, p = 0.91, ges < 0.01; spindle frequency during TMR periods: BF_10_ = 0.53, F_(1)_ = 0.36, p = 0.55, ges = 0.01; group by spindle frequency: BF_10_ = 0.32, F_(1)_ = 0.005, p = 0.95, ges < 0.01) (see Figure 2.B for details).

### CONSORT 2010 checklist of information to include when reporting a randomised trial

| Section/Topic | Item No | Checklist item | Reported on page No |
| --- | --- | --- | --- |
| **Title and abstract** | 1a | Identification as a randomised trial in the title | 1 |
|  | 1b | Structured summary of trial design, methods, results, and conclusions (for specific guidance see CONSORT for abstracts) | 2 |
| **Introduction** | | | |
| Background and objectives | 2a | Scientific background and explanation of rationale | 3-5 |
|  | 2b | Specific objectives or hypotheses | 4-5 |
| **Methods** | | | |
| Trial design | 3a | Description of trial design (such as parallel, factorial) including allocation ratio | 7, 20-21 |
|  | 3b | Important changes to methods after trial commencement (such as eligibility criteria), with reasons | NA |
| Participants | 4a | Eligibility criteria for participants | 20 |
|  | 4b | Settings and locations where the data were collected | 20 |
| Interventions | 5 | The interventions for each group with sufficient details to allow replication, including how and when they were actually administered | 20-23 |
| Outcomes | 6a | Completely defined pre-specified primary and secondary outcome measures, including how and when they were assessed | 23-28 |
|  | 6b | Any changes to trial outcomes after the trial commenced, with reasons | Supplementary Material  14-15 |
| Sample size | 7a | How sample size was determined |  |
|  | 7b | When applicable, explanation of any interim analyses and stopping guidelines | 20 |
| Randomisation:  Sequence generation | 8a | Method used to generate the random allocation sequence | 21 |
|  | 8b | Type of randomisation; details of any restriction (such as blocking and block size) | 21 |
| Allocation concealment mechanism | 9 | Mechanism used to implement the random allocation sequence (such as sequentially numbered containers), describing any steps taken to conceal the sequence until interventions were assigned | 21 |
| Implementation | 10 | Who generated the random allocation sequence, who enrolled participants, and who assigned participants to interventions | 21 |
| Blinding | 11a | If done, who was blinded after assignment to interventions (for example, participants, care providers, those assessing outcomes) and how | 21 |
|  | 11b | If relevant, description of the similarity of interventions | 22-23 |
| Statistical methods | 12a | Statistical methods used to compare groups for primary and secondary outcomes | 23-24, 27-28 |
|  | 12b | Methods for additional analyses, such as subgroup analyses and adjusted analyses | 25-28 |
| **Results** | | | |
| Participant flow (a diagram is strongly recommended) | 13a | For each group, the numbers of participants who were randomly assigned, received intended treatment, and were analysed for the primary outcome | NA |
|  | 13b | For each group, losses and exclusions after randomisation, together with reasons | 27 |
| Recruitment | 14a | Dates defining the periods of recruitment and follow-up | 20 |
|  | 14b | Why the trial ended or was stopped | 20 |
| Baseline data | 15 | A table showing baseline demographic and clinical characteristics for each group | 5-6 |
| Numbers analysed | 16 | For each group, number of participants (denominator) included in each analysis and whether the analysis was by original assigned groups | 23 |
| Outcomes and estimation | 17a | For each primary and secondary outcome, results for each group, and the estimated effect size and its precision (such as 95% confidence interval) | 7-16  &  Supplementary Material  1-9 |
|  | 17b | For binary outcomes, presentation of both absolute and relative effect sizes is recommended | NA |
| Ancillary analyses | 18 | Results of any other analyses performed, including subgroup analyses and adjusted analyses, distinguishing pre-specified from exploratory | 10-16  Supplementary Material  6-9 |
| Harms | 19 | All important harms or unintended effects in each group (for specific guidance see CONSORT for harms) | 20 |
| **Discussion** | | | |
| Limitations | 20 | Trial limitations, addressing sources of potential bias, imprecision, and, if relevant, multiplicity of analyses | 19 |
| Generalisability | 21 | Generalisability (external validity, applicability) of the trial findings | NA |
| Interpretation | 22 | Interpretation consistent with results, balancing benefits and harms, and considering other relevant evidence | 16-19 |
| **Other information** | | | |
| Registration | 23 | Registration number and name of trial registry | 20 |
| Protocol | 24 | Where the full trial protocol can be accessed, if available | 20 |
| Funding | 25 | Sources of funding and other support (such as supply of drugs), role of funders | 28 |

*We strongly recommend reading this statement in conjunction with the CONSORT 2010 Explanation and Elaboration for important clarifications on all the items. If relevant, we also recommend reading CONSORT extensions for cluster randomised trials, non-inferiority and equivalence trials, non-pharmacological treatments, herbal interventions, and pragmatic trials. Additional extensions are forthcoming: for those and for up to date references relevant to this checklist, see [www.consort-statement.org](http://www.consort-statement.org).

### List of Deviations from Pre-registration

|  | Preregistered | Final report |
| --- | --- | --- |
| 1 | Experiment 2, SRT single task, Offline consolidation:  Change in performance index (PI) between the first 2 blocks immediately after the nap+TMR intervention (post-nap) and the last 2 blocks of learning immediately prior to the intervention. | Change in **reaction time** between the first 2 blocks immediately after the nap+TMR intervention (**first 2 blocks of learning**) and the last 2 blocks of **test** immediately prior to the intervention.  Moreover, this analysis was joint together with that of retention (see 2 for further explanation). |
|  | **Justification**: As shown in the study by Nicolas et al. (Nicolas et al., 2024), the accuracy did not change with practice, and therefore RT appeared as a clearer proxy for motor learning and consolidation.  *Note: although in the preregistered statistical analysis plan we write 4 blocks, we need to account that they alternate between sequences. For this reason in this table we always report the notation* ***2 blocks****.* | |
| 2 | Experiment 2, SRT single task, Retention:  Change in **PI** between the first 2 blocks after the 24-hour retention period (post-night) and the last 2 blocks of Retest 1 immediately after the 2-hour NAP+TMR intervention. | Change in **reaction time** between the first 2 blocks after the 24-hour retention period (**first 2 blocks of learning**) and the last 2 blocks of **test pre-nap**. |
|  | **Justification**: Additionally, we chose to use the test pre-nap as a baseline for both the offline consolidation and the retention, in order to allow for comparisons between the two relative changes in the statistical analysis, as done in other studies (Nicolas et al., 2024) | |
| 3 | The primary outcome will be analysed with a repeated measures Analysis of Variance (ANOVA) with the following factors and conditions: 2 blocks * 2 sessions (pre-nap/post-nap) * 2 interventions (TMR/no TMR) * 2 groups (PD/HOA) | The primary outcome was analysed with a **Bayesian ANOVA**, complemented with a frequentist ANOVA, with the following factors: group (PD/HOA), intervention (TMR/no TMR), **Time (post-nap/post-night changes relative to pre-nap),** and their interactions as fixed effects, and subject as random effect. |
|  | **Justification:** To provide a clearer overview of learning across sessions, we opted for joining the Offline consolidation and Retention analysis with one model. This increased the power for highlighting possible differences. We tested for possible differences between blocks when calculating the relative changes post-nap and post-night, as we did not include the factor block as preregistered.  Also, we opted for reporting Bayesian statistical results (i.e., Bayes Factors) as they can provide a clearer interpretation of the effects given the underpowered nature of this study. | |
| 4 | Experiment 2 - SRT dual tasking: Offline consolidation:  Difference in PI between sequences A and B assessed across the 2 blocks of dual tasking immediately after the 2-hour NAP+TMR intervention (post-nap).  Experiment 2 - SRT dual tasking: Retention  Difference in PI between sequences A and B assessed across the 2 blocks of dual tasking after the 24-hour retention period (post-night). | Difference in **dual-task costs measured with reaction time** between the reactivated and non-reactivated sequence assessed across the 2 blocks of dual tasking immediately after the 2-hour NAP+TMR intervention (post-nap).  Difference in **dual-task costs measured with reaction time** between the reactivated and non-reactivated sequence assessed across the 2 blocks of dual tasking after the 24-hour retention period (post-night). |
|  | **Justification**: see (1) for explanation on the choice of reaction time.  We opted for using the dual-task costs (relative difference between dual-task and single-task performance) as a proxy of automaticity as it can more clearly show the impairment in automaticity typically seen in PD. | |
|  | Analyses reported under **Dual tasking performance (PI)** pertaining experiment 2 (points 2, 4, 5) and analyses reported under **Dual task cost (ΔPI)** (point 4, 5) in the statistical analysis plan were not performed given the underpowered nature of the study and the high granularity of the analyses. | |
